# Loss of salt iodization harmed child survival and academic achievement in Ethiopia

**DOI:** 10.64898/2026.07.08.26357562

**Authors:** Robel Alemu, Kibrom Tafere, Dawd Gashu, Edward J. M. Joy, Elizabeth H. Bailey, R. Murray Lark, Martin R. Broadley, William A. Masters

## Abstract

The introduction of salt iodization is associated with improved health and socioeconomic outcomes, but is not yet universally adopted and not always sustained. Using a quasi-experimental event study with difference-in-differences over space and time, we quantify the impacts of iodine deficiency in utero and infancy on childhood mortality and later academic achievement in Ethiopia, comparing cohorts born just before and after the May 1998 border closure that interrupted access to iodized salt. Rural children with fewer months of early-life exposure to iodized salt scored lower on standardized secondary-school exams, especially in districts with low environmental iodine, with excess deaths emerging in infancy and persisting through early childhood. These findings reveal the long-term benefits of salt iodization for health and education, especially for people with low intake of iodine from environmental sources.

## Main

Salt iodization is endorsed by the World Health Organization as a safe, cost-effective, and scalable intervention to prevent iodine deficiency^1^, yet is not universally implemented and sustained over time^2–4^. Randomized controlled trials in such diverse settings as Ethiopia^5^, New Zealand^6^, and Albania^7^ have demonstrated that iodine supplementation before pregnancy and during early childhood significantly enhances cognitive performance, but identifying and quantifying the long-term benefits of salt iodization at population scale is challenging due in part to the difficulty of measuring intake and confounding factors that coincide with iodization programs^4,8^. Historical evidence from the United States^9–12^ and Switzerland^13^ links early life iodization to better educational attainment and socioeconomic outcomes, while more recent data addresses salt iodization and schooling in Denmark^14^, China^15,16^ and India^17^, as well as linking the use of iodized oil capsules to school completion and employment status in Tanzania^18,19^. Even when iodization is introduced it is not always sustained, due in part to limited evidence of its value when the population no longer experiences visible symptoms of severe iodine deficiency such as goiter.^20^

The sudden withdrawal of iodized salt from Ethiopia in May 1998 provides a unique setting in which to isolate the effects of iodine deficiency on health and education. The country was an early adopter of universal salt iodization, prompted by high observed deficiency rates resulting from low levels of naturally occurring iodine and use of artisanal salt mined from inland deposits with negligible iodine content^21–23^. By the early 1990s, nearly universal iodized salt use was achieved through imports from a processing facility by the sea in Eritrea^24,25^. However, in May 1998 the Ethiopia-Eritrea conflict closed that border and abruptly halted access to iodized salt. In response, the Ethiopian government lifted its 1996 ban on the sale of non-iodized salt, leading to widespread reversion to low-iodine sources. By 2005 iodized salt usage had plummeted to 4%^22,24,26^, and the national micronutrient survey reported a median urinary iodine level of 24.5 µg/L, consistent with severe iodine deficiency, alongside goiter rates as high as 40% in school-aged children and 36% of women of reproductive age^21^. That prompted a gradual rise in iodized salt use and a return to mandatory iodization in 2012^27^, reaching 85% coverage by 2015^21,28^. This interruption and the subsequent necessary reliance on local food systems creates a large-scale natural experiment allowing us to investigate the consequence of iodine deficiency.

This study offers several distinct contributions to the literature by analyzing this context. First, we identify plausibly causal impacts by examining an exogenous shock – the abrupt and prolonged disruption of an established iodization program – rather than the gradual rollout typically studied. Second, we employ a methodological approach that utilizes high-resolution environmental data on soil iodine to measure exposure, improving upon prior studies that relied on coarse baseline goiter rates. Third, in contrast to work focusing largely on educational attainment (years of schooling), we examine long-term impacts on educational achievement (standardized test scores on a wide range of subjects), alongside effects on mortality. We hypothesize that the withdrawal of iodized salt would be most detrimental to children in locations with naturally low iodine in local soils, as rural populations in these areas rely heavily on local subsistence agriculture and lack alternative dietary iodine sources^29–31^.

To test this hypothesis, we employ dose-response difference-in-difference (DID) and event-study empirical frameworks, utilizing variation in naturally occurring iodine across three major agricultural regions (Amhara, Oromia and Tigray) which together account for nearly 70% of Ethiopia’s population and nearly 90% of its cultivated grain crop area^32^. We estimate the effect of iodine exposure in utero and infancy on two main outcomes: (i) student scores on Ethiopia’s nationally standardized secondary school exam (2003-2019), and (ii) child survival using Demographic and Health Surveys (2000 and 2005)^26,33^. We validate the pathway from environmental to biological iodine status by linking soil, grain and urinary iodine concentration data from the Ethiopian Micronutrient Survey (2015)^34^. Our identification relies on comparing cohorts born one-, two-, and three-year windows before and after the 1998 border closure, capturing variation in the timing of exposure during key developmental periods relative to the local soil iodine endowment.

We check the robustness of our results using a set of falsification tests and complementary analyses designed to rule out alternative explanations. First, we explicitly address the potential confounding role of the Ethiopia-Eritrea war by controlling for district-level conflict intensity and fatalities, ensuring that our results capture the nutritional shock rather than direct exposure to violence. Second, we further validate our findings by: (i) testing for effects using spatial variation in selenium that were not disrupted in 1998, testing the biological specificity of the iodine channel; (ii) examining anthropometric outcomes (height and weight) sensitive to general nutritional factors but not specifically to iodine; and (iii) analyzing urban children near Addis Ababa, whose food supply is predominantly sourced from regional markets and therefore likely unaffected by local soil nutrient profiles. These robustness checks would reject the expected null effect if our findings were caused by factors other than the hypothesized role of iodized salt in health and cognitive development.

## Results

### Timing of introduction, withdrawal and return of iodized salt in Ethiopia

Nationally representative surveys conducted between 1995 and 2015 reveal the high level and then sudden withdrawal followed by gradual recovery in household use of iodized salt in Ethiopia^28,33–36^. After its introduction, coverage in 1995 reached 80% of households reporting the use of iodized salt, all of which was imported from Eritrea^34^. Following the outbreak of war and closure of the Ethio-Eritrean border in May 1998, access to new iodized salt supplies was disrupted; although remaining household, shop, or distribution-chain stocks may have persisted briefly, available sources do not quantify how long such stocks lasted. We therefore interpret the 1998 border closure as marking the onset of rapid deterioration in iodized salt access, rather than the instantaneous disappearance of iodized salt from all households. Household use of any iodized salt declined quickly to 28% of households by 2000 and just 4% by 2005 (Figure 1A)^35^. Coverage remained limited through 2011 (15%), but increased to 44% by 2014, and rose sharply to 90% by 2015 after the reinstatement of mandatory iodization in 2012 (Figure 1A)^34^. This introduction, collapse and recovery of iodized salt access, captured across six national surveys, provides the basis for our empirical strategy leveraging regional and temporal variation in early-life exposure to iodine deficiency.

**Figure 1.**
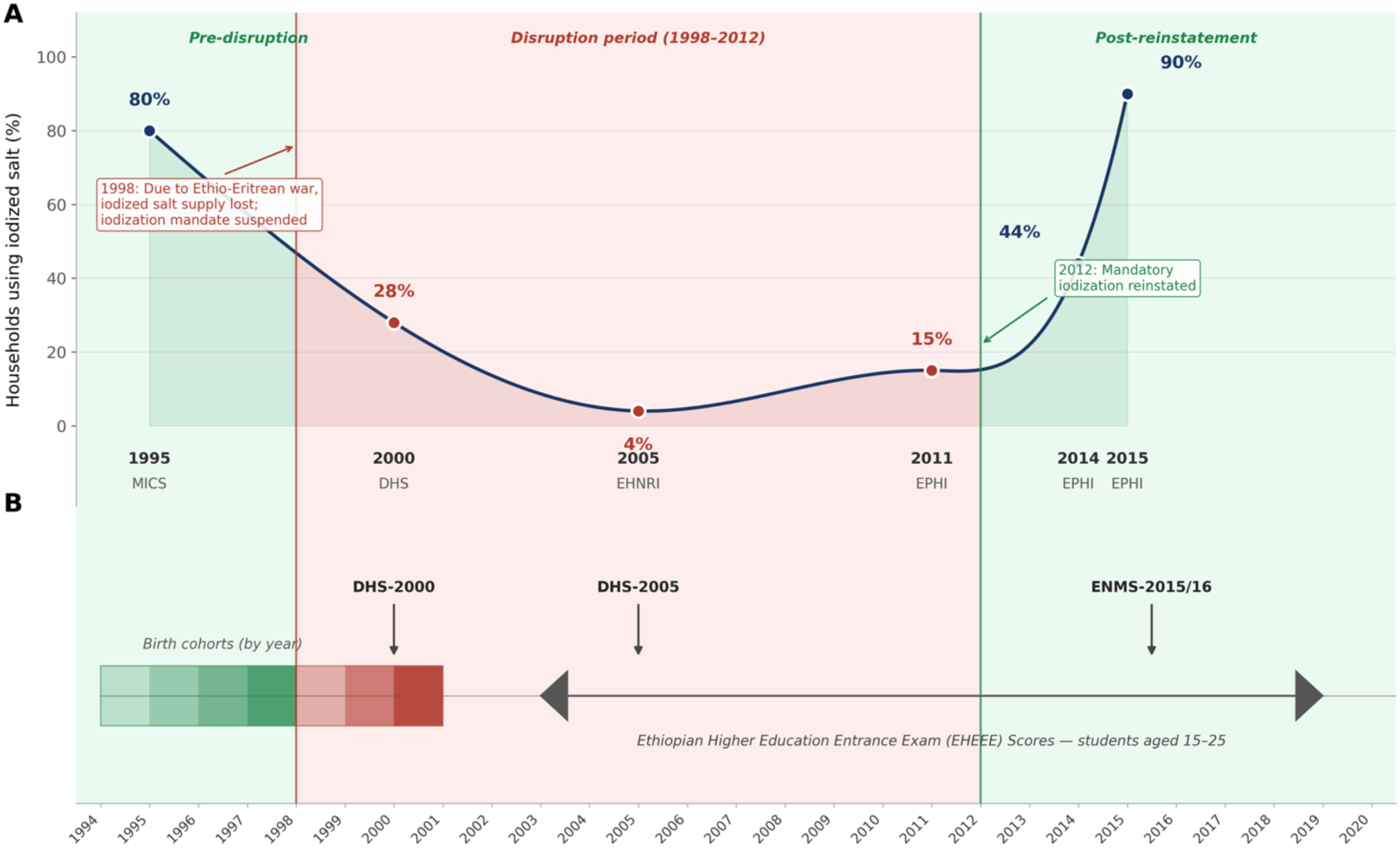
Iodized Salt Access and Timeline of Exposure and Measurement in Ethiopia. *Notes:* (A) Percentage of Ethiopian households reporting use of any iodized salt, based on nationally representative surveys: MICS (Multiple Indicator Cluster Survey), DHS (Demographic and Health Survey), EHNRI (Ethiopian Health and Nutrition Research Institute), EPHI (Ethiopian Public Health Institute), and ENMS (Ethiopian National Micronutrient Survey). The smooth line connects survey estimates. Background shading delineates three policy phases: pre-disruption (green, before 1998), disruption period (red, 1998–2012), and post-reinstatement (green, after 2012). The red vertical line marks 1998, when the Ethio-Eritrean war led to the loss of iodized salt supply and suspension of Ethiopia’s national iodization mandate. The green vertical line marks 2012, when mandatory iodization was reinstated. (B) Timeline of key events and birth cohorts affected by the 1998–2012 iodine disruption. Gradient shading within the cohort band reflects estimated exposure to iodine deficiency: darker red indicates more-affected post-war birth cohorts (1998–2000); darker green indicates less-affected pre-war cohorts (1994–1997). Also shown are nationally representative survey rounds used to measure household iodized salt coverage (DHS-2000, DHS-2005, ENMS-2015/16) and the period of Ethiopian Higher Education Entrance Exam (EHEEE) score data available for analysis (2003–2019, students aged 15–25). Soil and grain nutrient samples were collected during the late-2017 and late-2018 harvest seasons; as noted in the Methods, soil iodine concentrations are stable over decadal timescales, supporting their use as proxies for environmental iodine availability during the disruption period.

### Study population and estimation samples

Our analysis draws on data from over one million rural individuals across Ethiopia’s three largest agricultural regions. Educational outcomes were assessed for 1,080,685 young adults who sat for the Ethiopian Higher Education Entrance Examination (EHEEE) between 2003 and 2019. Estimation samples vary by birth cohort window: 232,526 individuals in the 1-year window, 452,136 in the 2-year window, and 603,569 in the 3-year window (Figure S1 and S8). Child survival and physical growth outcomes were evaluated using data from 8,185 rural children surveyed in the 2000 and 2005 rounds of the Ethiopian Demographic and Health Survey (DHS)^33,37^, with sample derivation summarized in Figure S9. Descriptive characteristics of the study cohorts across datasets are reported in Table S1.

### Mapping spatial variation in naturally occurring iodine and selenium

Identifying the effects of iodized salt in this study is made possible by data on naturally occurring iodine and selenium concentrations in soils and staple grains. Using geospatial sampling and statistical modeling, 1,352 soil samples were collected across Amhara, Oromia, and Tigray to estimate average nutrient concentration levels for all 385 administrative districts. Iodine was measured as the organically bound fraction extracted with 10% tetramethylammonium hydroxide (TMAH), and selenium as the soluble fraction extracted using 0.01 M potassium nitrate (KNO₃). Mean district-level soil iodine concentration was 10.18 mg/kg (range: 1.19–23.05), while selenium averaged 1.85 µg/kg (range: 0.009–5.15) (Figure 2, Table S1). Iodine levels were highest in western and parts of eastern Oromia, declined across Amhara from west to east, and were generally low in Tigray (Figure 2A). Selenium exhibited a distinct spatial pattern, shaped by underlying geological variation (Figure 2B).

**Figure 2.**
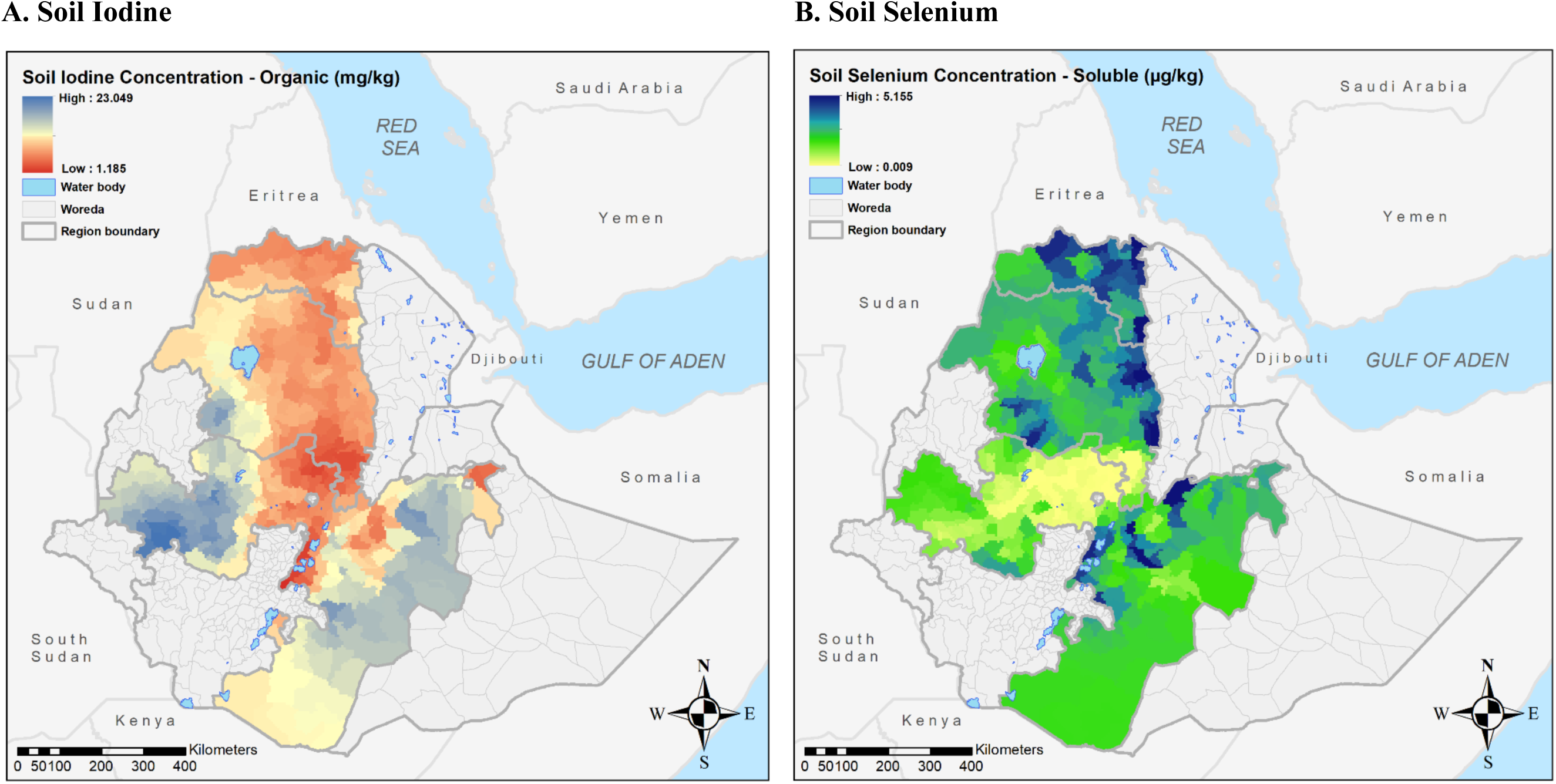
Spatial distribution of district-level average soil iodine and selenium concentrations in Ethiopia. *Notes*: The map illustrates the spatial distribution of soil nutrients across all districts in Ethiopia’s Amhara, Oromia, and Tigray regions. Panel A shows the average concentration of organic soil iodine, representing the “organically bound fraction” extracted using 10% tetramethyl ammonium hydroxide (TMAH). Panel B depicts the average concentration of soluble soil selenium, representing the “soluble” fraction extracted using 0.01 M KNO₃. The data were estimated using methods described in Gashu et al. (2021) and summarized in the supplemental information. District-level nutrient estimates were generated using methods adapted from Gashu et al. (2021) and the conditional simulation procedure described in Supplementary Note A3. The maps were created by the authors in ArcGIS using district boundary shapefiles and predicted district-level nutrient estimates; no previously published maps or third-party cartographic artwork were reproduced or adapted.

### Linking soil iodine to dietary intake and human iodine status

Our identification strategy relies on the premise that naturally occurring soil iodine influences iodine concentrations in locally grown crops and, indirectly, in animal-source foods^38,39^. This premise is especially plausible in rural Ethiopia, where household diets are tightly coupled to local agricultural conditions. Evidence from the 2011 and 2016 Ethiopian LSMS-ISA and Household Consumption and Expenditure Surveys shows that rural households obtain roughly 70–80% of staple grain consumption from own production or local markets, with limited reliance on long-distance food sourcing^29–31^. These localized food systems also imply limited exposure to industrially processed or imported foods made with iodized salt, reducing the likelihood that such foods substantially decouple iodine intake from local environmental conditions in rural Ethiopia^30,40,41^. In the absence of widespread supplementation or food fortification, spatial variation in soil iodine is therefore expected to translate directly into variation in dietary iodine intake when iodized salt is unavailable or inconsistently used. This soil → crop → diet pathway is well documented in agronomic and nutritional studies, including recent geospatial analyses linking soil iodine to crop iodine and population iodine status across sub-Saharan Africa^38,39,42,43^, and mirrors established evidence for selenium in Ethiopia^44^ and Malawi^45^. Groundwater may also contribute to naturally occurring iodine exposure^46,47^; because soil iodine and groundwater iodine are shaped by shared local geochemical and hydrological conditions^48–50^, this pathway would likely reinforce, rather than undermine, the use of soil iodine as a proxy for broader environmental iodine availability.

We provide direct empirical support for this mechanism using two complementary analyses. First, we show a strong relationship between soil and grain iodine concentration (Table S3). A 1% increase in organic soil iodine is associated with a 0.12% increase in grain iodine (p < 0.01), even after adjusting for topographic and climatic conditions. Because organic soil iodine is geochemically stable over long horizons, it captures persistent differences in the iodine content of locally produced foods rather than short-run agronomic or seasonal fluctuations^51,52^. Second, we demonstrate that a 1% increase in average grain iodine is associated with a 1.15% increase in urinary iodine concentration among school-aged children (p < 0.01), with consistent estimates across alternative model specifications (Table S4). This association remains significant even after adjusting for household consumption of adequately iodized salt, although the point estimate is modestly attenuated. Although collected after the period studied here, these nationally representative biomarker data provide external validation that districts with higher soil and grain iodine also exhibit higher biological iodine status in children

Together, these findings support the use of soil iodine as a meaningful proxy for long-term dietary iodine exposure in environments where households depend heavily on locally produced food and where fortification coverage is incomplete or disrupted. We emphasize that soil and grain iodine are not intended to measure individual intake precisely, but rather to capture persistent environmental differences in iodine availability that become salient when iodized salt access collapses. Because spatial groundwater iodine data were not available for the study regions, we could not incorporate this exposure pathway directly. Similarly, iodine-containing fortified blended foods and ready-to-use supplementary foods may be important iodine sources in food aid–dependent populations, but available evidence from Ethiopia indicates that such products were primarily delivered through targeted nutrition interventions rather than consumed broadly across the rural populations central to our analysis^53,54^. To the extent that groundwater, food aid, migration, dietary heterogeneity, or interpolation uncertainty weaken the link between soil iodine and individual consumption, such mismatch would attenuate estimated effects rather than generate spurious dose–response relationships, implying that our estimates are conservative lower bounds of the true impacts of early-life iodine deficiency.

### Early life disruption to iodized salt lowered exam performance in iodine-deficient areas

Table 1 presents our main results, showing that the loss of access to iodized salt in 1998 significantly reduced performance on secondary school exams of students born around that time, particularly in districts with low levels of naturally occurring iodine. In this difference-in-differences framework with continuous treatment, we compare exam scores between children born just before and just after the disruption, and quantify how the impact varied by the soil iodine concentration of their district. Children in rural areas born just after the loss of iodized salt scored 0.02 to 0.04 standard deviations lower on the national university entrance exam than those who were born just before, when their mothers had access to iodized salt (p < 0.01), and children born in districts with higher natural iodine levels experienced significantly smaller losses. For every 1 SD increase in district-level soil iodine, test scores among post-disruption cohorts were 0.03 to 0.09 SD higher (p < 0.01), indicating a protective effect of local dietary iodine. These results are robust across cohort comparisons and strongest in the 3-year window, which captures more sustained exposure during critical developmental periods in utero and early childhood. To put the magnitude of this effect in concrete terms, among children born in the three-year window following the disruption, those residing in 1 SD lower soil iodine districts scored approximately 8.3 points lower than their pre-disruption counterparts in districts with average iodine levels, based on a pre-disruption standard deviation of 63.9 points.

**Table 1.** Dose-response of secondary-school exam scores to local soil iodine for children born after loss of access to iodized salt.

|  |  | **Rural Districts** |  |  | **Urban Districts around Addis Ababa** |  |  |
| --- | --- | --- | --- | --- | --- | --- | --- |
|  |  | **(treatment effect, due to use of locally grown food)** |  |  | **(falsification test, due to use of food from elsewhere)** |  |  |
|  |  | Born 1998<br>vs 1997 | Born 1998-99<br>vs 1996-97 | Born 1998-2000<br>vs 1995-97 | Born 1998<br>vs 1997 | Born 1998-99<br>vs 1996-97 | Born 1998-2000<br>vs 1995-97 |
| Post |  | -0.024*** | -0.037*** | -0.041*** | -0.075 | -0.237 | -0.279 |
|  | SE | (0.009) | (0.013) | (0.015) | (0.012) | (0.213) | (0.247) |
|  | 95% CI | (-0.042, -0.006) | (-0.063, -0.01) | (-0.071, -0.012) | (-0.294, 0.146) | (-0.655, 0.181) | (-0.762, 0.205) |
|  | P value | 0.008 | 0.007 | 0.006 | 0.528 | 0.296 | 0.285 |
| Post × Local Iodine |  | 0.025** | 0.08*** | 0.089*** | -0.049 | -0.241 | -0.297 |
|  | SE | (0.011) | (0.015) | (0.018) | (0.099) | (0.188) | (0.232) |
|  | 95% CI | (0.004, 0.047) | (0.051, 0.109) | (0.054, 0.123) | (-0.243, 0.146) | (-0.61, 0.128) | (-0.752, 0.157) |
|  | P value | 0.023 | 0.000 | 0.000 | 0.635 | 0.232 | 0.228 |
| Pre-war Avg. Score |  | 361.610 | 360.014 | 356.407 | 340.954 | 336.749 | 333.973 |
|  | SD | (63.928) | (63.928) | (63.928) | (76.82) | (76.82) | (76.82) |
| N |  | 232,393 | 451,877 | 603,208 | 5,171 | 9,7794 | 13,735 |
| R<sup>2</sup> |  | 0.281 | 0.265 | 0.262 | 0.434 | 0.398 | 0.381 |

*Notes:* Data shown are estimated coefficients (standard errors) from the dose-response DID model described in Equation (2). The dependent variable for all columns is the child's 3-subject cumulative score in EHEEE's cumulative score (7-subjects), expressed as z scores. The bottom row of the table shows the mean and standard deviation of their raw scores to show magnitudes (for example, an effect size of roughly 0.05 implies a difference of about 3 points around the average score of about 356 points). Results for urban districts surrounding Addis Ababa (Oromia Special Zone) are a placebo test, because food supplies to the city bring grain from elsewhere. Each column refers to cohorts born 1, 2, or 3 years after or before the loss of iodized salt in 1998 due to the closure of the Ethiopia-Eritrea border, where the indicator variable 'Post' equals 1 for children born in or after the calendar year 1998, and 0 otherwise. 'Local Iodine' is the z-score of district-level average soil iodine concentration. All models adjust for sex, academic stream, and include test year, school and district fixed effects. Standard errors, robust and clustered at the district level, are shown in parentheses. Significance levels are \*\*\*p<0.01, \*\*p<0.05, \*p<0.1.

We complement our difference-in-differences design with an event-study analysis that traces how the relationship between soil iodine and exam scores evolved across annual birth cohorts (Figure 3). Coefficients represent the marginal effect of soil iodine on test scores for each cohort, relative to the reference year 1997—the last cohort with full early-life exposure to iodized salt. Among earlier cohorts born between 1994 and 1996, the relationship between soil iodine and test performance is slightly negative and relatively stable, suggesting no divergence in trends prior to the disruption. This pattern supports the plausibility of the parallel trends assumption required for causal identification. Beginning in 1998, however, the soil iodine–score gradient shifts sharply upward and becomes increasingly positive for cohorts born in 1999 and 2000, before stabilizing. This progression indicates the growing impact of iodine deficiency over time and highlights the protective role of naturally occurring iodine as reliance on iodized salt diminished.

**Figure 3.**
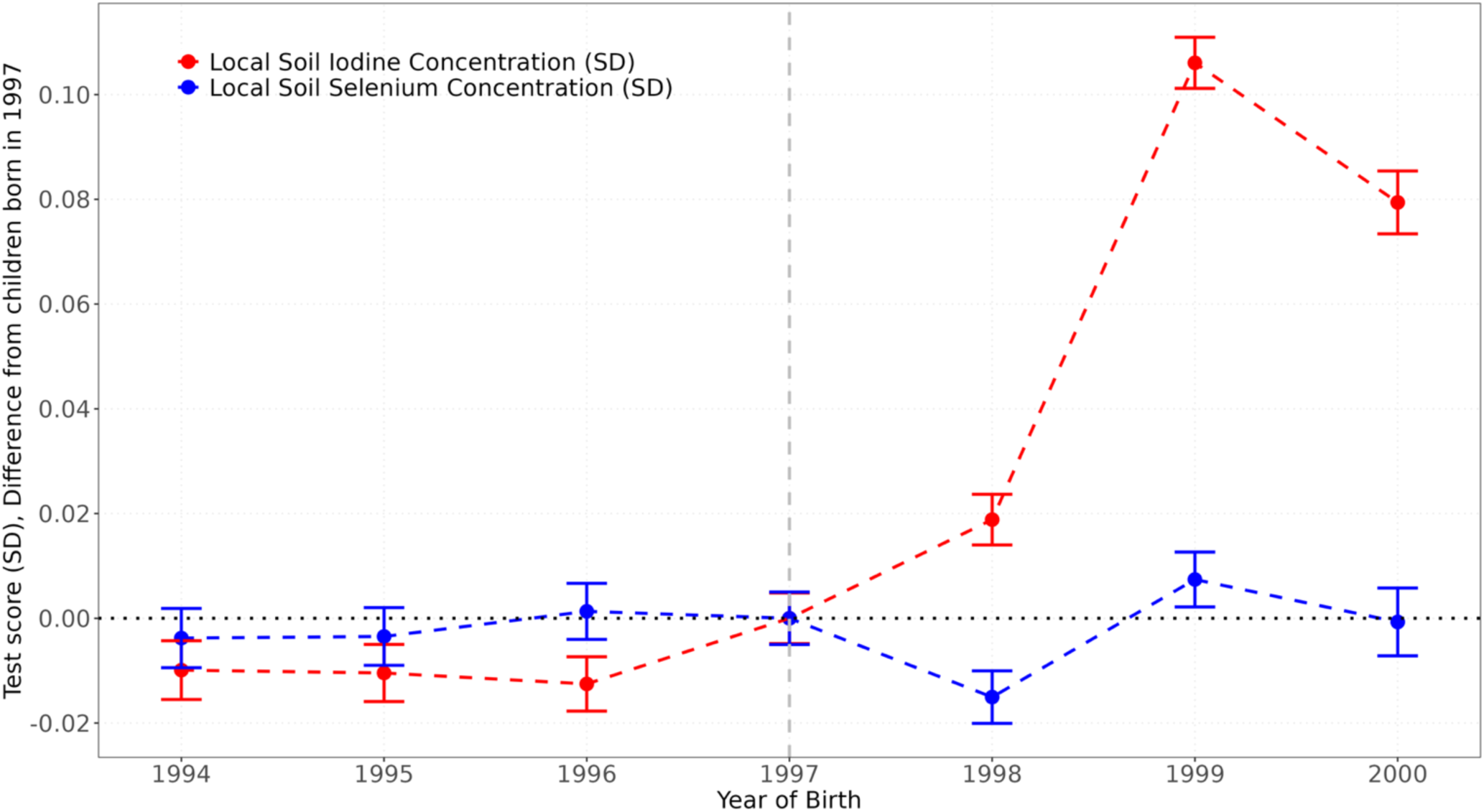
Event study of dose-response in exam scores to 1 SD higher local soil nutrients by year of birth. *Notes*: Data shown are estimated regression coefficients (points) with 95% confidence intervals based on heteroskedasticity-robust standard errors clustered at the district (woreda) level. Statistical significance was assessed using two-sided tests and no adjustment was made for multiple comparisons. The unit of analysis is the individual student exam record (N = 684,163) from 385 rural districts/woredas; no technical or biological replicates were used. The Y-axis shows the effect sizes for a child’s 7-subject cumulative score in the EHEEE, expressed as standard deviations. Red dots and confidence intervals represent the relationship between soil iodine levels and test scores, while blue dots and confidence intervals depict the falsification results for soil selenium. The falsification analysis tests the robustness of findings, as selenium levels are not expected to be influenced by the iodized salt disruption. Coefficients for each birth year are shown relative to the reference year 1997, meaning the results capture how test scores for children born in each year differ from those born in 1997 in response to a marginally higher (1SD) soil iodine concentration relative to the mean student in 1997. The chart reflects how the dose-response of soil iodine to test scores changes for children born before, during, and after the iodized salt disruption of 1998. Results are limited to rural districts in the three study regions and account for academic stream, test year, and district fixed effects.

We also examine heterogeneity in the iodine effects across test subjects and by sex. Estimated effects differ across subjects, with relatively larger protective effects observed for some subjects than others, but without a clear or uniform pattern across cognitive domains (Figures S2–S3). We additionally find slightly larger protective effects of marginal soil iodine for girls than for boys, although these differences are not statistically significant (Figure S4). Such variation may reflect differences in how early-life iodine deficiency affects specific components of cognitive development rather than broad, uniform impacts on test performance. The direction of the gender pattern is consistent with prior randomized trials and nutritional studies documenting greater vulnerability of female fetuses to early-life iodine deficiency^18,55,56^.

We then assess the robustness of these findings through a series of complementary analyses designed to rule out alternative explanations. A key concern is that the withdrawal of iodized salt coincided with the Ethiopia–Eritrea conflict, which could have independently affected child health and educational outcomes^57^. Although the baseline models include district and test-year fixed effects, these do not fully capture heterogeneity in conflict exposure across districts and birth cohorts. To address this, we re-estimate both the dose–response and event-study models after explicitly controlling for district-year conflict exposure using Armed Conflict Location and Event Data (ACLED) data^58^ on total conflict events and conflict-related fatalities (Supplementary Note A4C). Incorporating these controls leaves the magnitude and statistical significance of the iodine interaction estimates essentially unchanged in both frameworks (Table S6), indicating that the observed iodine gradient is not driven by residual war-related shocks.

We further probe the biological specificity of the iodine channel through multiple falsification tests, described in detail in the Methods. First, replacing soil iodine with soil selenium – an essential micronutrient that is similarly transmitted from soils to plants and people but, unlike iodine, was unaffected by the 1998 disruption and not subject to fortification – yields no association with exam performance in either the DID or event-study specifications, ruling out the possibility that our findings reflect micronutrient variation in general rather than iodine specifically (Table S5; Figure 3). The sole exception is a marginally significant coefficient (p < 0.1) in the broadest 3-year birth-cohort window; however, this result does not recur in narrower windows. Second, applying the same model to urban districts surrounding Addis Ababa, where food supplies are largely sourced from outside local agricultural systems and thus unlikely to reflect district-level soil iodine variation, produces similarly null results, suggesting our findings are not artifacts of the data sources or model structure (Table 1; Figure S5). Third, substituting anthropometric outcomes — birthweight, height-for-age, and weight-for-height — for exam scores and survival also yields uniformly null results across all cohort windows and soil iodine specifications, indicating that our main findings are not driven by broader nutritional or health shocks that would be expected to affect physical growth (Table S10; Figure S7). Taken together, the robustness of our estimates to conflict controls and the null results across all three falsification tests strengthen the interpretation that the observed effects reflect the disruption of iodized salt rather than confounding factors or artifacts of the study design.

For a further assessment of the dose–response relationship—and to address concerns that cognitive benefits from iodine may plateau or reverse at high exposure levels—we first replace the continuous soil iodine measure with terciles of the district-level iodine distribution (Figure 4). Test scores increase monotonically across terciles, consistent with a smooth and approximately linear gradient rather than discrete thresholds or reversals. We complement this descriptive check with parametric robustness tests designed to capture potential departures from linearity and to assess whether extreme iodine exposure yields attenuation or adverse effects. Specifically, we augment the baseline specification with a quadratic interaction between post-disruption birth cohorts and soil iodine to allow for curvature in the exposure–response function (Table S7), and we conduct an upper-tail robustness check by interacting post-disruption exposure with an indicator for districts in the top decile of the iodine distribution (Table S8). Across cohort windows, these extensions yield estimates that are highly consistent with the linear specification, indicating that the main dose–response results are not driven by functional-form assumptions or by unusually high iodine exposure. Details on the implementation of these nonlinearity checks are provided in Supplementary Note A4B.

**Figure 4.**
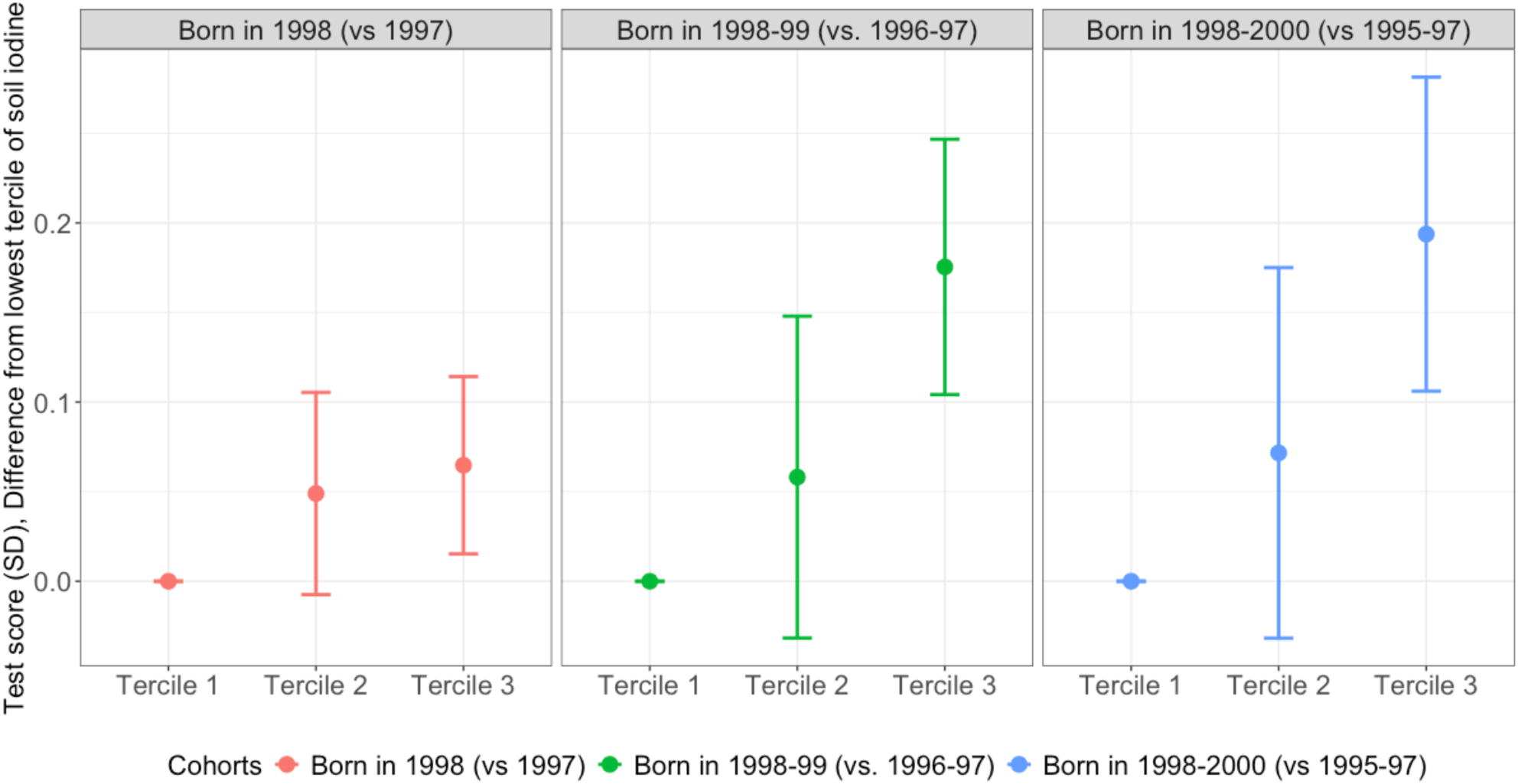
Categorical dose-response in exam scores to terciles of local soil iodine by birth cohort. *Notes*: Data shown are estimated coefficients and 95% confidence intervals based on standard errors clustered at the district (woreda) level. The unit of analysis is the individual student exam record; no technical or biological replicates were used. Sample sizes were N = 232,392 student exam records from 383 districts/woredas for the one-year comparison (1998 vs. 1997), N = 451,873 from 384 districts/woredas for the two-year comparison (1998–1999 vs. 1996–1997), and N = 603,204 from 385 districts/woredas for the three-year comparison (1998–2000 vs. 1995–1997). Statistical significance was assessed using two-sided tests, and no adjustment was made for multiple comparisons. The Y axis shows effect sizes for the child’s 3-subject cumulative score in English, Aptitude, and Civics from the EHEEE, expressed as z scores, associated with the interaction of having a medium or high level of district-average soil iodine (relative to the lowest tercile) with being born just after the loss of iodized salt, relative to the USI period before 1998. Colors refer to the birth-year cohorts born within one (red), two (green), and three (blue) years of the cutoff. Results are restricted to rural districts of the three study regions. All models control for academic stream and include fixed effects for test year, and district.

We further assess robustness to uncertainty in the geostatistical prediction of district-level soil iodine by implementing a bootstrap procedure that jointly propagates uncertainty in environmental exposure and exam-score sampling variability (Supplementary Note A4A). Because district iodine measures are conditionally simulated using observed individual observations, treating them as fixed regressors could understate total uncertainty if prediction noise meaningfully affects the estimates. To address this concern, we resample districts with replacement—drawing repeatedly from the empirical distribution of simulated iodine levels—and resample exam takers within districts, preserving the structure of the quasi-experimental design. Re-estimating the full specification in Equation (2) across 500 bootstrap replications yields an empirical distribution of the treatment and dose–response coefficients that closely aligns with the main estimates, with comparable magnitudes and confidence intervals (Table S9). These results confirm that our findings are robust to uncertainty in soil-iodine prediction as well as to sampling variation in student outcomes.

### Salt iodization improved child survival but not physical growth

We examine child survival using Kaplan-Meier curves based on births reported in the 2000 and 2005 DHS surveys (Figure 5). Mortality was substantially higher among children born after 1998 in districts with below median (lower-half) of soil iodine, with elevated risk beginning in the first month and persisting through the first two years of life. Cumulative mortality in these areas approached 25-30%, compared to less than 10% elsewhere. While high in absolute terms, these levels are consistent with published estimates for rural Ethiopia in the late 1990s and early 2000s—particularly in remote, drought-affected, and conflict-adjacent areas^27,59–61^. In contrast, differences by soil selenium were small and not statistically meaningful. Importantly, the soil-iodine mortality gradient remains evident after accounting for local conflict exposure, indicating that these patterns are not driven solely by contemporaneous violence.

**Figure 5.**
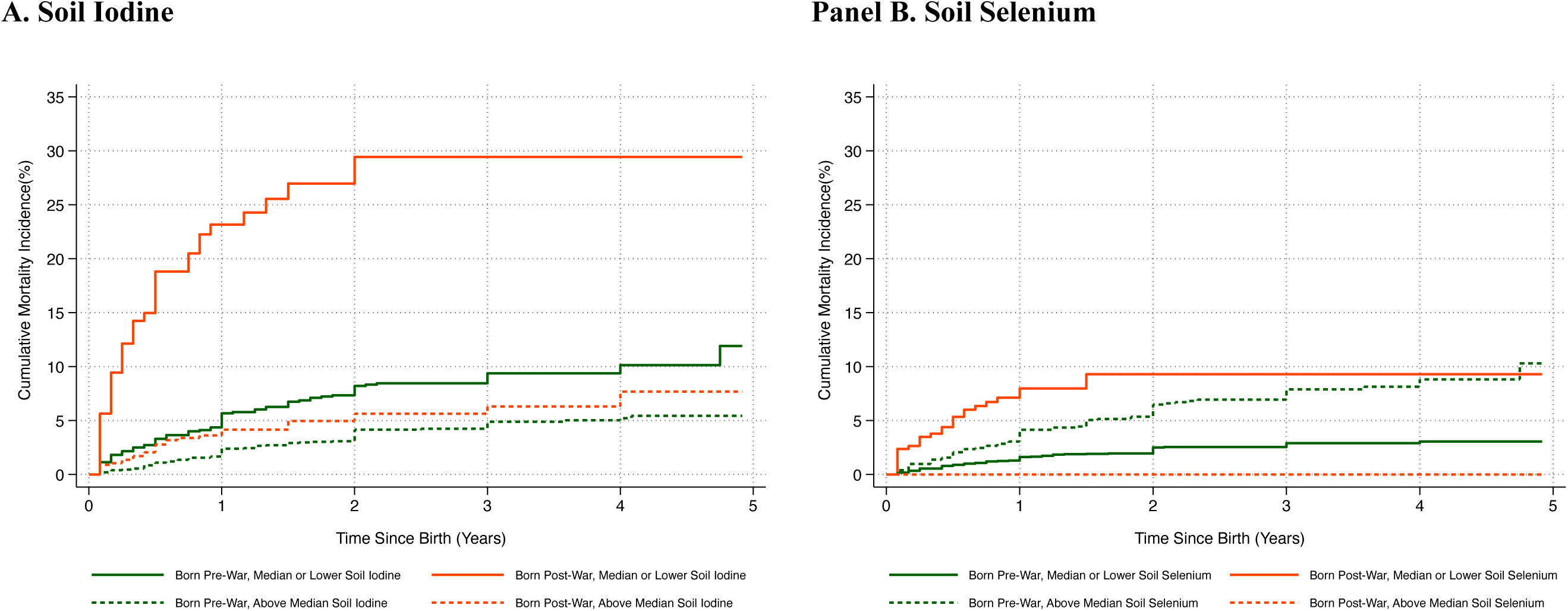
Cumulative mortality of under-five year rural children by birth cohort and level of soil nutrients. *Notes*: This figure displays Kaplan-Meier cumulative incidence curves for mortality, stratified by children’s birth cohort relative to the start of the war (3 years before and after 1998), and district-level soil iodine and selenium concentrations. At baseline (time=0, the child’s birth), all children enter the risk set with a survival probability of 1 (100%). Each subsequent year, the survival probability adjusts according to S_(t+1)=S_t×(N_(t+1)-D_(t+1))/N_(t+1), and the cumulative failure probability as 1-S_t. The curves are generated using the “sts graph” command in Stata MP, and is adjusted for child sex, maternal education, maternal BMI, maternal age at first birth, child’s birth order, area of residence, wealth index, birth year, and district fixed effects. Panel A illustrates the treatment effects: solid lines for children from the lowest tercile of soil iodine districts, and short-dashed lines for those from the highest tercile, with pre-war cohorts in dark green and post-war in dark orange. Panel B depicts the effects for soil selenium, which is known to play a role in general health but was not remedied by salt iodization prior to the war, with solid lines for children from the lowest quartile of soil selenium districts, and short-dashed lines for those from the highest quartile, with pre-war cohorts in dark green and post-war in orange-red. Results are restricted to students who resided in rural districts of the three study regions.

The availability of DHS data allows us to use anthropometric indicators in a falsification test, checking whether our main results are actually an artifact of study design caused by factors that would affect physical growth, in contrast to iodine’s effects on cognition and mortality. Across all cohort comparisons, we find no significant dose-response effects on birthweight, height-for-age, or weight-for-age (Table S10 and Figure S7). Although limited sample size reduces statistical power for these outcomes, the absence of detectable effects on physical growth—contrasted with pronounced effects on survival and later cognitive performance—is consistent with iodine deficiency operating primarily through neurological development and early-life survival rather than through generalized growth impairment.

## Discussion

We examined the long-term consequences of the introduction, withdrawal and then later return of iodized salt on Ethiopian child survival and later academic achievement. Leveraging data on spatial variation in naturally occurring soil iodine, we found that putative early-life iodine deficiency significantly reduced secondary school performance and increased early-childhood mortality among rural children. Children born when they and their mothers lacked access to iodized salt scored lower on national university entrance exams (0.02–0.04 SD), with larger deficits in areas with lower environmental iodine. These patterns are evident across multiple birth-cohort windows and remain stable across alternative specifications of iodine exposure.

Our results reveal that iodine deficiency in utero and infancy has lasting consequences for survival and exam scores that can be avoided by use of iodized salt, with significant dose response to naturally occurring iodine in maternal and child diets. Event-study estimates for exam scores reinforce this result, showing that the protective influence of soil iodine grew with each birth cohort after 1998, peaking among those born in 2000. The absence of differential pre-trends in the soil iodine–exam score gradient prior to the disruption supports the identifying assumptions of the difference-in-differences design. These results underscore the importance of iodine for fetal development and early childhood and demonstrate the long-term consequences of even temporary interruptions to iodized salt access.

Our findings also reveal substantial mortality effects. Rural infants born when their household had less access to iodized salt faced significantly higher risk of death in the first two years of life, particularly in districts with low naturally occurring iodine. Although high in absolute terms, these mortality levels are consistent with independent estimates for rural Ethiopia during the late 1990s and early 2000s, especially in remote, drought-affected, and conflict-adjacent areas^62–66^. These patterns are also consistent with evidence from smaller clinical and programmatic studies: a randomized trial in Indonesia reported a 72% reduction in infant mortality following iodized salt supplementation^67^, while an intervention in China using iodine-enriched irrigation reduced infant mortality by more than one-third^68^. Together, these findings indicate that iodine deficiency can have large survival consequences in low-iodine environments, particularly during early life^49,50^.

The exam-score and mortality findings should be interpreted jointly. Because exam outcomes are observed only among children who survived early childhood, remained in school, and sat for the national entrance examination, the academic estimates necessarily reflect a selected subset of the exposed birth cohorts. The elevated mortality in low-iodine districts suggests that children most severely affected by early-life iodine deprivation were less likely to be represented in the exam data; additional selection through delayed grade progression or school dropout would further attenuate observed academic effects. Thus, the estimated test-score differences likely represent lower bounds of the full population-level impact of early-life iodine deprivation.

The magnitude of these effects is consistent with the severity of iodine deficiency in Ethiopia as a low-iodine environment. Data from the 2015/16 ENMS, collected after iodization had largely recovered and when approximately 85–90% of households were using iodized salt, show that school-aged children had a median urinary iodine concentration of just 105–108 µg/L, barely above the WHO sufficiency threshold, while women of reproductive age remained deficient at 86–97 µg/L (Table S2). This implies that without iodized salt, moderate to severe iodine deficiency would be the norm across much of rural Ethiopia, given that locally grown crops and water are insufficient to meet requirements. Consistent with this, district- and regional-level studies conducted during and after the disruption period documented goiter prevalences of 37–55% among schoolchildren across multiple regions, including Amhara, Oromia, Tigray, and Southwest Ethiopia^69–72^. The 1998–2012 withdrawal of iodized salt therefore removed what was, in many rural districts, the only meaningful dietary iodine source, making the observed effects on child cognition and survival biologically plausible and contextually expected.

Placed in the context of prior work, our findings contribute to a growing literature on the long-term consequences of iodine supplementation while addressing a distinct and policy-relevant margin. Existing large-scale studies have primarily examined the introduction or expansion of iodization programs, often using baseline indicators of iodine deficiency to study later schooling attainment or enrollment outcomes^15,17,73–75^. In contrast, our setting allows us to study the consequences of a prolonged collapse in iodized salt access, followed by recovery, and to examine how this temporal shock interacted with exogenous spatial variation in naturally occurring iodine. This focus on policy interruption rather than rollout speaks directly to the stability of fortification gains—an issue of practical importance in low-income settings where coverage is uneven and vulnerable to disruption. Methodologically, combining a time-varying shock with contemporary geospatial measures of iodine availability provide a complementary perspective to approaches based on historical goiter prevalence or deficiency rates. Substantively, by examining standardized, high-stakes exam performance and child survival at population scale, we document lasting human capital losses that arise when iodization systems fail, even temporarily.

Across outcomes, our findings are robust to a wide range of potential confounders and alternative explanations. Results are unchanged when accounting for spatial and temporal heterogeneity in conflict exposure, when testing for nonlinearity in the iodine dose–response relationship using quadratic and upper-tail specifications, and when propagating uncertainty in geostatistical iodine prediction through a joint district-level bootstrap. Additional falsification tests—using soil selenium as a placebo exposure, anthropometric outcomes such as birthweight, height and weight that are sensitive to general health and nutrition but not directly to iodine, and urban populations less reliant on local food systems—consistently yield null effects. Taken together, these complementary analyses substantially narrow the set of plausible mechanisms that could generate the observed patterns absent iodine deficiency, and indicate that the results are not driven by functional-form assumptions, extreme exposure values, conflict-related shocks, or broader nutritional changes.

Several limitations warrant consideration. First, because our design is quasi-experimental rather than randomized, residual confounding cannot be fully ruled out—especially if other shocks around 1998 shifted discretely and correlated with environmental iodine in ways not captured by the available controls. Second, we do not observe individuals longitudinally, so we cannot directly track early-life iodine exposure, migration, household food sourcing, or school continuation; relatedly, survivorship and selective progression to the exam may attenuate exam-score impacts, so the estimates for achievement are likely conservative. The administrative exam data also contain limited information on student or parental background beyond sex, academic stream, school, district, birth cohort, and exam year, limiting our ability to characterize who progressed to exam taking or how selection changed over time. Third, iodine exposure is proxied using contemporary geospatial soil and grain iodine rather than contemporaneous biomarkers during the disruption period, and the district exposure measures are predicted with interpolation error; while our validation exercises and bootstrap procedures reduce these concerns, measurement error remains.

We also could not directly incorporate groundwater iodine, which may contribute to environmental iodine exposure in some Ethiopian settings, because district-level groundwater iodine data were not available for the study regions. Future work combining prospective biomarker data (maternal and child urinary iodine), richer dietary and market-integration measures, and linked administrative schooling records would help pinpoint critical windows of vulnerability, quantify selection into exam taking, and more directly map policy coverage to true iodine intake. Overall, while Ethiopia’s unique history and geography enabled our analysis, the dose-response nature of our findings suggests broader relevance – particularly for rural populations in regions where salt iodization programs are absent, inconsistent, or ineffective^76–80^.

In conclusion, our study shows that early-life iodine deficiency caused by low concentrations in locally grown food or water can not only affect human health as shown previously^81,82^, but also affect schooling outcomes among rural children in the general population. Even after salt iodization became national policy, temporary lapses had long-term consequences for rural communities reliant on locally sourced food. As many countries continue to struggle with achieving and sustaining adequate iodine coverage^76,77,79,83–85^, our findings highlight the importance of resilient fortification systems and demonstrate the long-term educational returns to micronutrient policy at scale. Our results combine prior work on environmental iodine variation^35,86,87^ with population-level evidence on health and schooling to document the enduring human capital costs of iodine deficiency among rural children in a low-income setting.

## Methods

This study uses data on iodine and selenium concentration in soils and cereal grains, matched with survey data on birth dates, mortality, and anthropometry in childhood, and also matched with birth dates and exam scores taken at the end of secondary school, across three major agricultural regions of Ethiopia (Amhara, Oromia, and Tigray). These three regions account for over 68 percent of the national population and nearly 90 percent of the country’s cultivated grain crop area^32^. We use difference-in-differences, and event study approaches to causal inference, focusing on the interaction between the duration of exposure to iodized salt in early life and variation in naturally occurring iodine in local foods. To control for artifactual results, we use three kinds of falsification tests: first with soil selenium (instead of iodine) because selenium is an essential trace element that plays an important role in thyroid function, but its availability was not affected by changes in salt iodization, then with urban districts (instead of rural) because children there would consume more food from elsewhere that is unaffected by their district’s soil iodine, and third with anthropometric outcomes (instead of mortality or test scores) because height and weight are known to be affected by many aspects of child health but have not been robustly associated with iodine deficiency.

### Ethics Statement

Grain and soil samples were collected from farmers’ fields and grain stores with informed consent from participating farmers. The GeoNutrition study protocols were reviewed by the Directors of Research at Addis Ababa University, who determined that ethical review was not required in Ethiopia. The study was also reviewed and approved in the United Kingdom by the University of Nottingham School of Sociology and the Social Policy Research Ethics Committee (BIO-1718-0004). Tufts University’s Institutional Review Board reviewed the protocols for the present study and determined that the research did not constitute human subjects research under DHHS and FDA regulations; consequently, full IRB review and approval were not required (IRB ID: STUDY00000776).

### Micronutrient content of soils and grain

Our study is based on the systematic sampling of soil and grain from representative sites, used with covariates for spatial prediction of district-level average iodine and selenium concentrations. The sampling frame was restricted to areas with a crop cultivation probability exceeding 90 percent and within 2.5 km of all-weather roads to ensure accessibility and increase the likelihood that food grown at that location would be consumed by people living nearby. The crop cultivation probability was derived from machine learning models applied to high-resolution satellite imagery and remote sensing covariates on a 250-m grid, details of which are provided in^87^. Samples were collected with the informed consent of households at the sampling site. In total, this approach yielded 1,352 soil and grain samples from 385 districts spanning the three study regions, as illustrated in Figure 2.

The soil and grain sampling procedure are summarized in the supplementary section A1 and prior literature ^86,88^. In brief, soil samples were analyzed for the three commonly reported fractions of soil iodine and selenium: soluble, adsorbed, and organic. These fractions are defined based on a sequential extraction process with different reagents, detailed in supplemental section A2. Analyses were performed at the University of Nottingham using an inductively coupled plasma mass spectrometer (ICP-MS). Individual measurements of each fraction from each study site were then used to estimate district average concentrations using conditional simulation as described in supplemental section A3, based on a geostatistical model of spatial dependence in soils around the sample sites. These district-level estimates serve as proxy measures of environmental exposure and capture the soil-to-grain nutrient pathway. Among the three fractions, we focused on organic iodine and soluble selenium as the primary exposure measures, because organic iodine typically constitutes most of the total soil iodine across diverse soil types, while soluble selenium (selenate) is the most readily bioavailable form for plant root uptake across soil systems.

### Validity of iodine in soils and grain as a predictor of human iodine status

Our study design rests on the transmission of naturally occurring iodine in soils to food and, thereby, to people who consume at least some locally grown food. In addition to our main results, we can test directly whether human iodine status is predicted by local iodine concentrations in soil and crops using the 2015/16 wave of Ethiopia’s National Micronutrient Survey (ENMS). The ENMS has a smaller sample size than DHS but includes data from more costly and invasive deficiency biomarkers. Of the 2,052 ENMS participants, a total of 945 had data for urinary iodine and also location data, placing them in districts for which we have estimated iodine concentrations in the soil and locally grown crops. In line with WHO criteria for school-aged children, iodine deficiency is defined as median urinary concentrations below 100 μg/L; thresholds differ for other population groups. Results for the pertinent ENMS variables in Table S2.

### Academic achievement in secondary school

Academic achievement at the upper-secondary level was assessed from students’ performance on the Ethiopian Higher Education Entrance Examination (EHEEE), a nationwide standardized exam administered at the end of grade 12. To capture scores for children born during and after Ethiopia’s first period of universal salt iodization in the 1990s, we compiled the country’s largest repeated cross-sectional database of all students who sat for the EHEEE from 2003 through 2019. A flow diagram in supplemental Figure S8 describes the exclusion criteria applied to reach our final estimation sample. To address potential privacy concerns, test score data was first de-identified by dropping all information other than birth dates, and we matched students to districts using GPS coordinates of the school the student attended when they sat for the EHEEE. Children are not expected to have been born or spent their early childhood at the location of their secondary school, but moves are more likely to have occurred within than between districts^89–91^. Studies suggest that most rural students remain near their place of birth during the critical early-life period when iodine exposure has the greatest impact, with relocations to urban centers for secondary schooling typically limited to nearby towns^90,91^. This minimizes the potential distortion of measured exposure to soil and grain iodine levels ^92,93^. Furthermore, to ensure that test scores are comparable across space and time, raw scores were standardized to z-scores by subtracting the mean from each student’s raw score and dividing by the standard deviation of scores for all students who were born in the same year, sat for the test in the same year, and resided in the same administrative region. This approach ensures that students in each district are compared only to peers of the same age who answered the same exam questions within the same administrative structure.

### Birthweight, child survival, and growth

Our child health data are from the 2000 and 2005 waves of Ethiopia’s Demographic and Health Surveys (DHS). The DHS employs a structured two-stage cluster design, selecting enumeration areas and then sampling households within each area. Our data are extracted from DHS birth recode files, resulting in one record for each child ever born to the respondent, typically the biological mother of the child (Figure S9). We then matched enumeration areas to districts for exposure to naturally occurring iodine in soils and food. Birth dates, birth weight where available, and whether the child was still alive at the time of the survey are recorded directly. Growth is computed as z scores of attained height and weight of children under five years of age, relative to the 2006 WHO reference population of healthy children at each age and sex, producing height-for-age (HAZ), weight-for-age (WAZ), and weight-for-height (WHZ) indicators calculated using the Stata command zscore06. Figure S9 shows the exclusion criteria for the final estimation sample.

### Study design and statistical analysis

This study compares outcomes for children born in time to benefit from Ethiopia’s initial adoption of salt iodization in the 1990s to outcomes for those born after access to iodized salt was cut off in May 1998, interacting with the amount of naturally occurring iodine available in their locally-grown foods. Each child’s year of birth is a discrete variable, while the child’s exposure to naturally occurring iodine is a continuous variable. Across birth years, the causal effect at dose *d* of natural iodine is 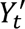 (*d*), the partial derivative of the difference in potential outcomes with respect to the dose, leading to the following average causal response (ACR) estimated over the entire sample^49^.

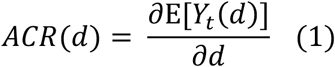

To estimate dose response, we control for observable characteristics of the child, and fixed effects for all time invariant attributes of their district or school, and all aspects of their survey or exam year, so as to focus on the interaction between the degree of naturally occurring iodine in their district and whether their birth timing exposed them to lack of iodized salt during critical periods of child development in utero and infancy:

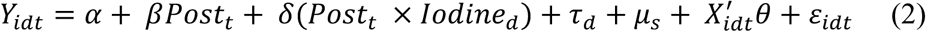

In equation (2), *Y*_idt_ denotes the outcome of interest for child *i*, born in year *t* and observed in district *d* with average soil iodine concentration *Iodine*_d_. *Post*_t_ is an indicator equal to 1 if child *i* was in the cohort born after withdrawal of iodized salt, *X*_idt_ represents a vector of individual-level covariates, *τ*_d_ are location fixed effects capturing all time-invariant characteristics of district or school *d* (including local iodine levels), *μ*_%_ is the survey-year or test-year fixed effects controlling for all common factors affecting outcomes measured in year *s,* and *ε*_idt_ is random error. The coefficient on the interaction term (*δ*) is the main parameter of interest and measures the difference in outcomes caused by naturally occurring iodine among those least protected by the presence of iodized salt. To facilitate interpretation all outcomes and the (*Iodine*) variable are standardized, reported in units of that variable’s standard deviation around its mean. Equation (2) does not contain a term for the independent effect of soil iodine on outcomes, as the district fixed effect absorbs all time-invariant district-level factors that affect our outcomes.

The child’s iodine status depends on maternal use of iodized salt or high-iodine foods before and during pregnancy and lactation, as well as the child’s own intake of iodized salt or high-iodine foods. In Ethiopia through the 1980s, non-iodized salt was mined using artisanal techniques from deposits in the Afar region. In the 1990s, there was a gradual rise in the use of iodized salt, especially after the 1996 ban on the sale of non-iodized salt for human use, until the border with Eritrea was closed and universal iodization policies were rescinded in 1998^94^. To isolate the effects of changing access to iodized salt from other trends or patterns, we compare cohorts born immediately before and after the cutoff using birth-year windows of three different sizes. The narrowest comparison uses two single-year cohorts, setting *Post*_t_ equal to 0 for children born in 1997 and 1 for those born in 1998. For longer exposure to iodized salt before the cutoff we compare two-year cohorts, setting *Post*_t_ equal to 0 for those born in 1996 or 1997 and 1 for births in 1998 or 1999. The largest cohort we consider had the most exposure to iodized salt, but also more variation in other conditions over time, setting *Post*_t_ equal to 0 for those born in 1995 through 1997 and 1 for births in 1998 through 2000. Each of the three cohort pairs compare groups of approximately equal size.

Event study specification offers an alternative to discrete cohorts, with successive comparisons between all children born in each year during and after salt iodization. The base year of peak iodization was 1997, so the event study regressions are specified as follows:

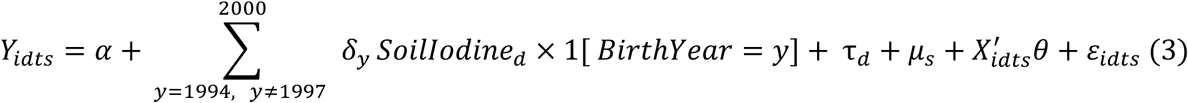

Our main coefficients of interest in Equation (3) are values for *δ*_y_, which measure the average causal response to district-level iodine soil levels for each birth year *y* from 1994 to 2000, with 1997 as the reference base year before the loss of iodized salt. Besides capturing change in access to iodized salt from year to year, the event study specification in Equation (3) allows testing for a difference before and after the 1998 cutoff. In this context, testing for pre-trends concerns whether the coefficients (*δ*_y_) are similar for children born in 1994 through 1997, and then diverge for those born in 1998 and later due to the cutoff in access to iodized salt. The coefficient *δ*_y_ is each birth- year’s cohort-specific impacts of a marginal increase in soil iodine content (1 SD) on outcome *Y*. All the other terms in Equation (3) are defined as in Equation (2).

Equations (2) and (3) are estimated using ordinary least squares (OLS) regression for continuous outcomes and linear probability regression models (LPM) for binary outcomes. In all regressions, we adjusted the standard errors of the estimated coefficients to account for potential spatial correlation by clustering at the district level. All continuous outcome variables and also district-average nutrient concentrations were transformed into standard deviations for ease of interpreting the effect sizes. The dose-response results from Equation (2) are presented in table form (Tables 1, Table S5-S10), while the event study results from Equation (3) are shown graphically (Figure 3).

### Testing for false positives via falsification tests

An important concern is whether our findings are false positives generated by the study design or how our data were collected and transformed for this study. We conducted three kinds of falsification tests to check whether our main results could be artifacts of the data and method.

First, we substituted soil selenium instead of iodine to determine whether our results might be driven by micronutrients in general rather than iodine. Selenium, like iodine, is an essential micronutrient for human health, including cognitive development. Both are transmitted from soils to plants and people. However, unlike iodine, selenium levels have remained stable over time, without supplementation or fortification programs analogous to salt iodization^95–97^. This makes soil selenium a suitable stability control for testing the specificity of our findings. If our results using soil iodine also held with soil selenium, we would know that the result was not actually driven by iodine in salt but by some other aspect of the study design and our data sources.

Second, we substituted urban instead of rural districts, focusing on the capital city (Addis Ababa) whose food supplies come primarily from elsewhere, in contrast to rural areas where mothers and children are likely to consume a larger share of their food from farms within their district^30,31,98^. This type of placebo test is potentially very sensitive to artifactual false positives because all of the data sources and model structure are the same, but the sample size is much smaller, so a null result could arise for that reason.

Third, we substituted physical growth, as measured by birthweight and the child’s attained height and weight, to test whether the observed improvements in survival and exam scores might be driven by broader factors such as diet quality, disease, or care practices, which are known to affect all of those outcomes^99,100^, in contrast to iodine whose effects are more specifically associated with thyroid function and cognition^56,82^. Like other falsification tests, if we had found significant results for physical growth, it would suggest that our results for survival and exam scores could be false positives caused by confounding factors or artifacts of our study design. Finding null results on all three falsification tests supports the validity of our causal inference, complementing our other robustness checks from alternative specifications and dose-response analysis.

## Data availability

Data on grain and soil iodine and selenium concentrations are available through the Rothamsted Research CKAN data repository (https://data.rothamsted.ac.uk/). The 2015/16 Ethiopian National Micronutrient Survey (ENMS), used to infer urinary iodine status in children and women of reproductive age, was provided by the Ethiopian Public Health Institute (https://ephi.gov.et) under a data-sharing agreement (FSND/011/2020). The 2000 and 2005 waves of the Ethiopia Demographic and Health Survey (DHS), used to analyze birthweight, child survival, and physical growth, were obtained from the DHS Program (https://dhsprogram.com/data/available-datasets.cfm) under the relevant data-use agreement. Data on students’ performance in the Ethiopian Higher Education Entrance Examination (EHEEE; 2003–2019) were obtained from the Education Assessment and Examination Service (EAES) of Ethiopia’s Ministry of Education and are available from the corresponding authorities subject to their data-sharing policies.

## Code availability

All code is freely available at https://github.com/robelalemu01/Impact-of-Iodized-Salt-Ethiopia.git.

## Data Availability

Data on grain and soil iodine and selenium concentrations are available through the Rothamsted Research CKAN data repository (https://data.rothamsted.ac.uk/). The 2015/16 Ethiopian National Micronutrient Survey (ENMS), used to infer urinary iodine status in children and women of reproductive age was provided by the Ethiopian Public Health Institute (https://ephi.gov.et) under a data-sharing agreement (FSND/011/2020). The 2000 and 2005 waves of the Ethiopia Demographic and Health Survey (DHS) used to analyze birthweight child survival and physical growth were obtained from the DHS Program (https://dhsprogram.com/data/available-datasets.cfm) under the relevant data-use agreement. Data on students performance in the Ethiopian Higher Education Entrance Examination (EHEEE; 2003 to 2019) were obtained from the Education Assessment and Examination Service (EAES) of Ethiopian Ministry of Education and are available from the corresponding authorities subject to their data-sharing policies.

## Acknowledgements

We thank the participating farmers and field sampling teams from the Amhara, Oromia, and Tigray Regional Bureaus of Agriculture, Ethiopia. The collection and analysis of grain and soil nutrient concentrations was led by Martin Broadley, together with Murray Lark, Edward Joy, Dawd Gashu, and Elizabeth Bailey. Mineral analytical support was provided by W. Broadley, S. Young, E. Bailey, L. Wilson, K. Davis, P. Muleya, S. Vasquez Reina, S. Dunham, J. Carter, and J. Hernandez. We thank Lucinda Toyama for research assistance; Andualem Mengistu for facilitating access to the educational achievement data; our dissertation committee members Cynthia Kinnan, Steven Block, and Lauren Schmitz; and Kyle Emerick, Margaret McMillan, David Cutler, and many other colleagues for their helpful feedback and suggestions on earlier versions of this work.

## Funding Statement

Robel Alemu’s work on this study was conducted as part of his doctoral dissertation at Tufts University and was funded by the Neubauer Family Foundation, with additional support from the Friedman School of Nutrition Science and Policy. This work was also supported, in part, through the Innovative Methods and Metrics for Agriculture and Nutrition Action (IMMANA) programme, led by the London School of Hygiene & Tropical Medicine (LSHTM), in partnership with Tufts University and the University of Sheffield. IMMANA is co-funded by UK Aid from the UK government and the Gates Foundation (INV-002962/OPP1211308). Additional support was provided through the GeoNutrition project, funded by the Gates Foundation (INV-009129) and the UKRI Biotechnology and Biological Sciences Research Council (BBSRC)/Global Challenges Research Fund (GCRF) (BB/P023126/1). Rothamsted Research receives strategic funding from the Biotechnology and Biological Sciences Research Council (BBSRC) and acknowledges support from the Growing Health Institute Strategic Programme (BB/X010953/1; BBS/E/RH/230003C). Under the grant conditions of the Gates Foundation, a Creative Commons Attribution 4.0 Generic License has been assigned to the Author Accepted Manuscript version that might arise from this submission.

## Author contributions

R.A. conceived the study, conducted all main analyses, and wrote the first draft of the manuscript. W.A.M. supervised the study design and execution, contributed to the interpretation of results, and revised subsequent manuscript drafts. K.T. co-supervised the project, facilitated data access, and provided critical input on the interpretation of findings. R.M.L. performed conditional simulations to predict district-level average soil and grain nutrient levels. E.H.B. led the laboratory analysis of the soil and grain nutrient concentrations. E.M.J. and M.R.B. led soil and crop studies underlying the analysis of naturally occurring iodine. All authors contributed to the writing, critically reviewed, and commented on the manuscript.

## Competing Interests

The authors declare no competing interests.

## Notes

### Competing Interest Statement

The authors have declared no competing interest.

