## Supplementary Information for "Loss of salt iodization harmed child survival and academic achievement in Ethiopia"

### Supplementary information for
### Loss of salt iodization harmed child survival
### and academic achievement in Ethiopia

#### Contents:

Figs. S1 to S9

Tables S1 to S10

Supplementary Notes

Supplementary References 1-20

Figure S1. Distribution of test-takers by birth-year and cohort

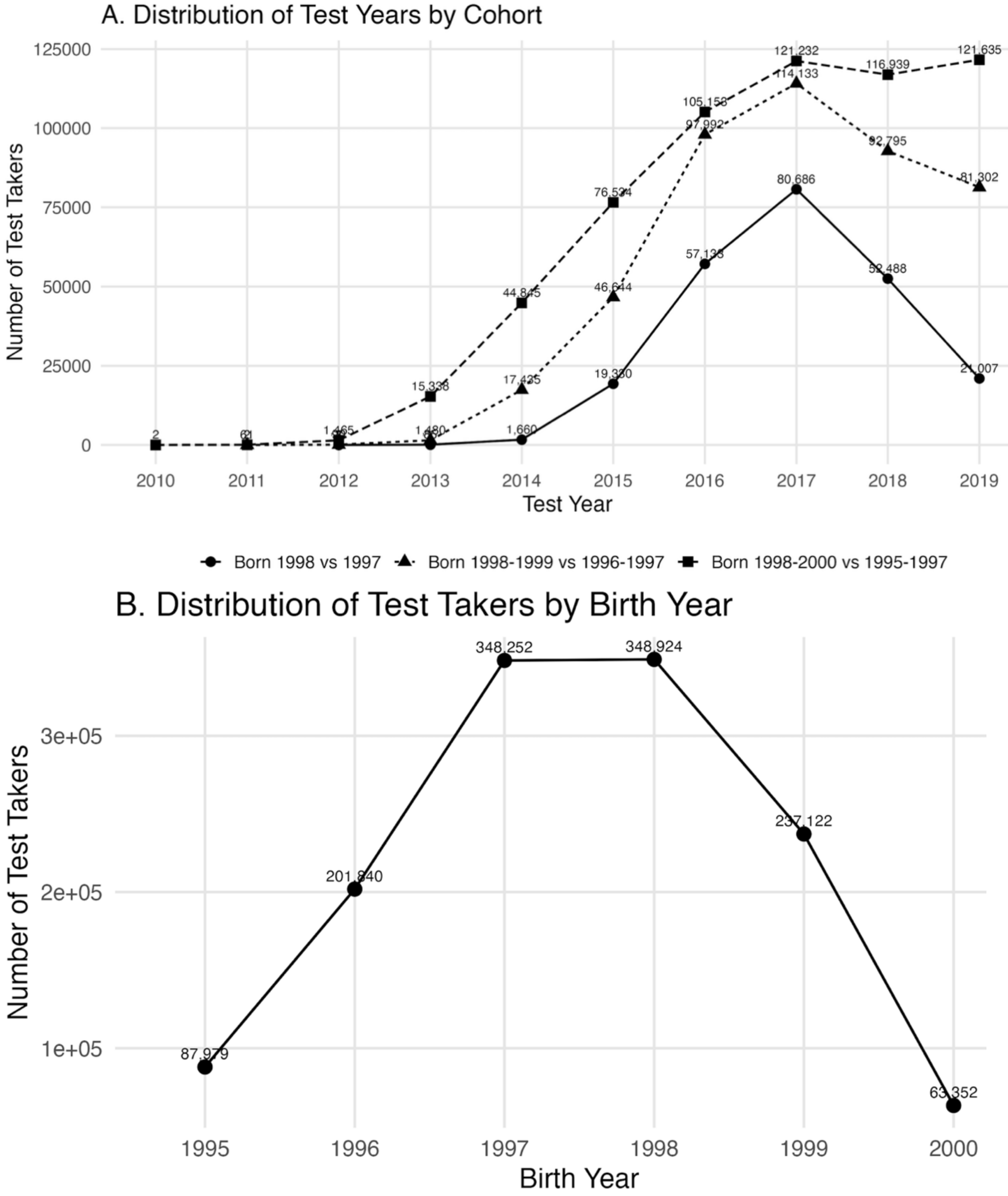

*Notes*: Panel A shows the number of individual student exam records by test year for the three birth-cohort comparison windows used in the difference-in-differences analysis: born 1998 vs 1997, born 1998–1999 vs 1996–1997, and born 1998–2000 vs 1995–1997. Panel B shows the number of individual student exam records by birth year across the cohort windows. Points show observed counts, with lines connecting adjacent years to aid visualization; no error bars or statistical tests are shown. The unit of analysis is the individual student exam record, and no technical or biological replicates were used. The full estimation sample used in the event-study specifications spans students born between 1988 and 2004 who sat for the EHEEE between 2003 and 2019 (N = 1,080,685). The cohort-restricted difference-in-differences samples comprise 232,526 student records for the one-year window, 452,136 for the two-year window, and 603,569 for the three-year window.

Figure S2. Differences in exam scores for students born just after (vs. before) the loss of iodized salt, by subject

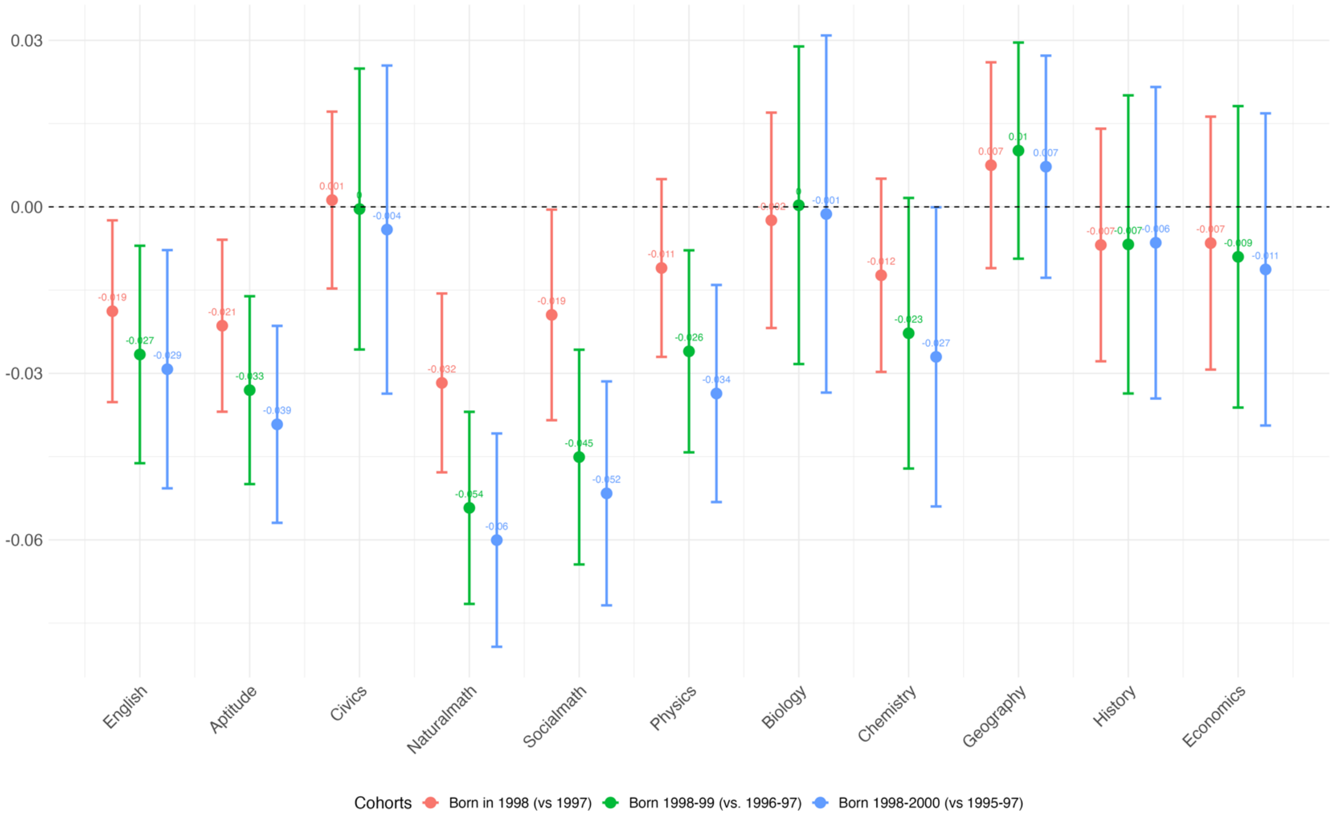

*Notes*: Data shown are estimated regression coefficients (points) with 95% confidence intervals calculated from heteroskedasticity-robust standard errors clustered at the district (woreda) level for the 'Post' term in the dose-response difference-in-differences (DID) model (Equation 2). Statistical significance was assessed using two-sided tests, and no adjustment was made for multiple comparisons. The unit of analysis is the individual student exam record; no technical or biological replicates were used. The coefficients represent the average difference in subject-specific test scores, measured in standard deviations, associated with being born in the one-year (red), two-year (green), or three-year (blue) post-disruption windows relative to students born the same number of years before the 1998 cutoff, independent of the protective effects of local soil iodine. The estimation sample is restricted to students residing in rural districts of the three study regions. All models control for academic stream where relevant and include fixed effects for test year, school, and district.

Figure S3. Dose response in exam scores to 1 SD higher soil iodine, for students born just after the loss of iodized salt, by subject

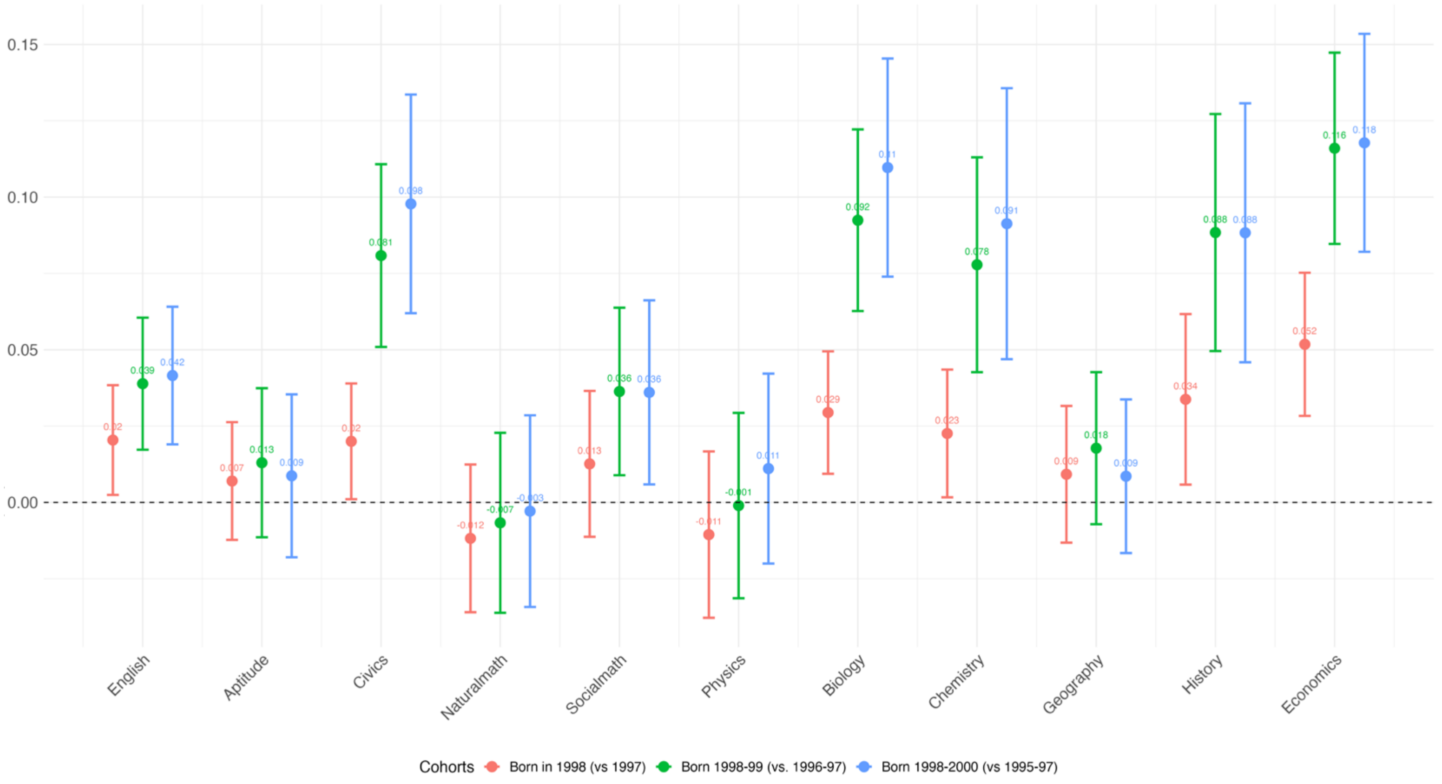

*Notes*: Data shown are estimated regression coefficients (points) with 95% confidence intervals calculated from heteroskedasticity-robust standard errors clustered at the district (woreda) level for the dose-response interaction term of the difference-in-differences (DID) model (Equation 2). Statistical significance was assessed using two-sided tests, and no adjustment was made for multiple comparisons. The coefficients represent the average difference in subject-specific test scores, measured in standard deviations, associated with a one standard deviation higher level of soil iodine in the child's district for students born in the one-year (red), two-year (green), or three-year (blue) post-disruption windows relative to students born the same number of years before the 1998 cutoff. The unit of analysis is the individual student exam record; no technical or biological replicates were used. Sample sizes were N = 232,392 student exam records from 383 districts/woredas for the one-year comparison, N = 451,873 from 384 districts/woredas for the two-year comparison, and N = 603,204 from 385 districts/woredas for the three-year comparison. The estimation sample is restricted to students residing in rural districts of the three study regions. All models control for academic stream where relevant and include fixed effects for test year, school, and district.

Figure S4. Dose response in exam scores to 1 SD higher soil iodine for males and females born just after the loss of iodized salt

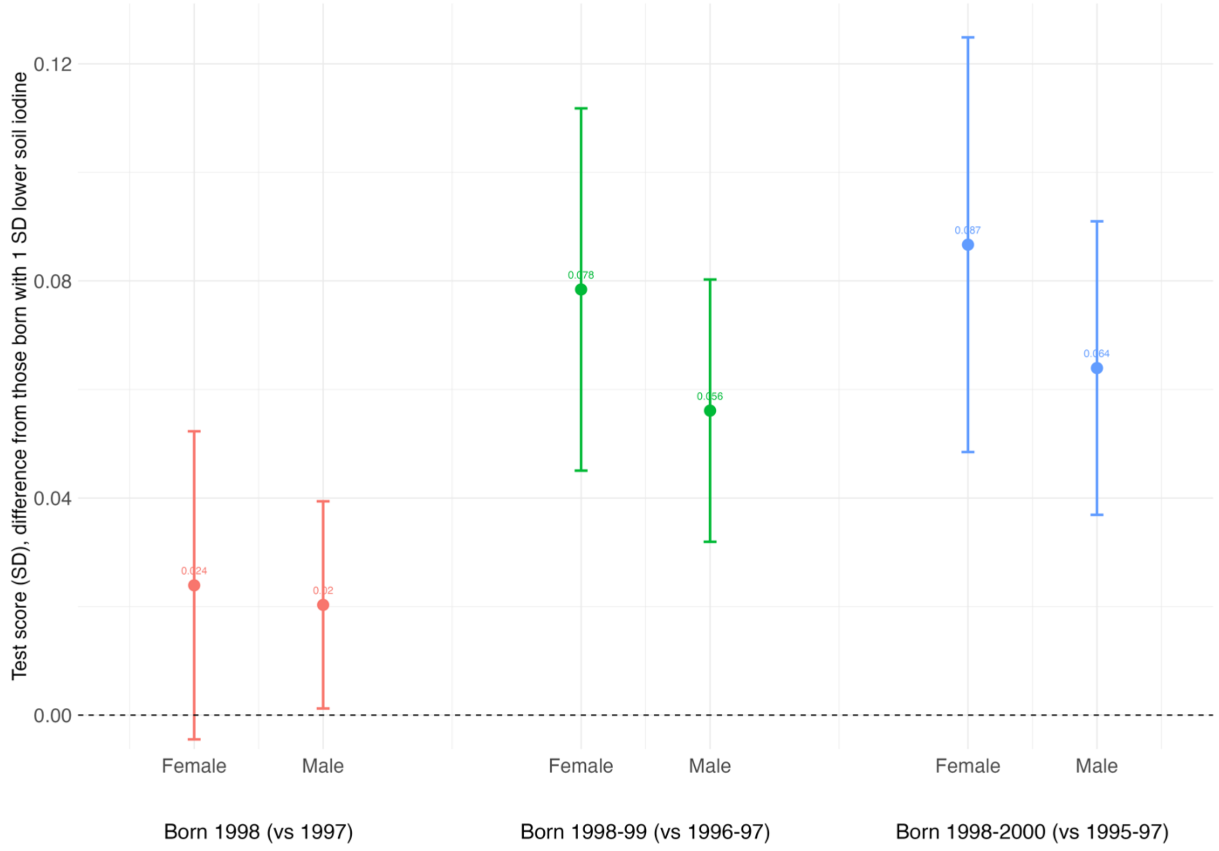

*Notes*: Data shown are estimated regression coefficients (points) with 95% confidence intervals calculated from heteroskedasticity-robust standard errors clustered at the district (woreda) level for the dose-response interaction term of the difference-in-differences (DID) model (Equation 2), with analyses stratified by sex. Statistical significance was assessed using two-sided tests, and no adjustment was made for multiple comparisons. The dependent variable is the student's standardized 3-subject cumulative score from the Ethiopian Higher Education Entrance Examination (EHEEE). The unit of analysis is the individual student exam record; no technical or biological replicates were used. Sample sizes were N = 232,392 student exam records from 383 districts/woredas for the one-year comparison, N = 451,873 from 384 districts/woredas for the two-year comparison, and N = 603,204 from 385 districts/woredas for the three-year comparison. All models control for academic stream and include fixed effects for test year and district.

Figure S5. Event study of dose response in exam scores to 1 SD higher soil nutrients, for students in urban (placebo) districts

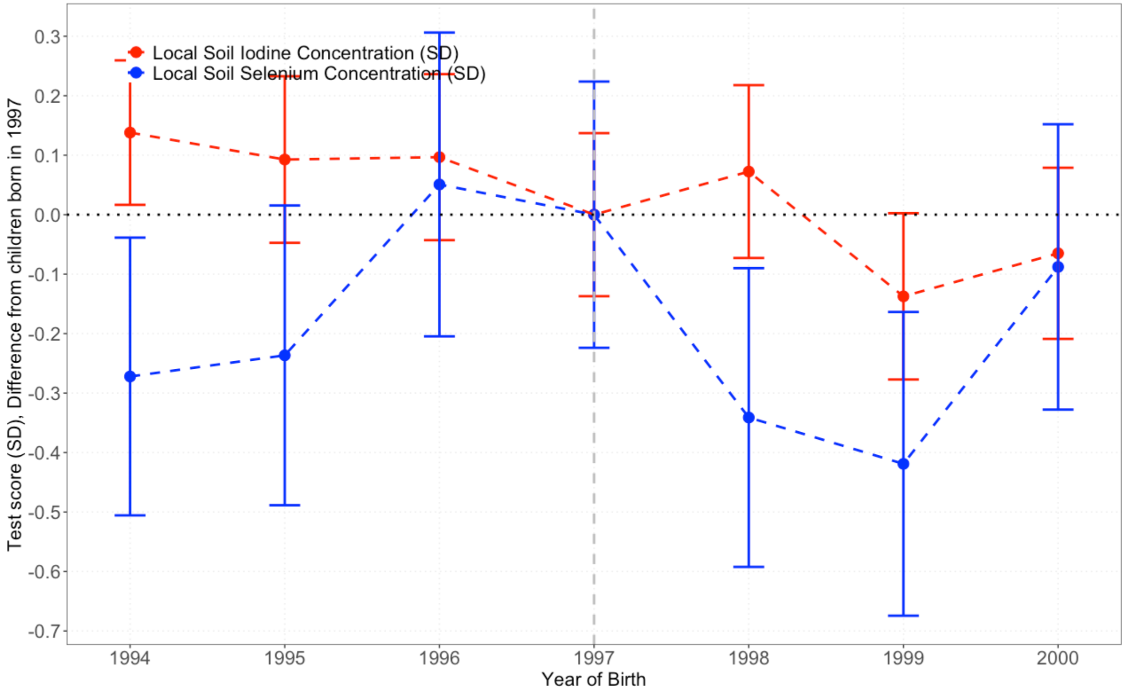

*Notes*: Data shown are estimated regression coefficients (points) with 95% confidence intervals calculated from heteroskedasticity-robust standard errors clustered at the district (woreda) level from the event-study dose-response analysis (Equation 3). Statistical significance was assessed using two-sided tests, and no adjustment was made for multiple comparisons. The unit of analysis is the individual student exam record (N = 9,604) from 7 urban districts/woredas; no technical or biological replicates were used. Year-specific sample sizes were: 1994, n = 1,040; 1995, n = 1,166; 1996, n = 1,323; 1997, n = 1,724; 1998, n = 1,593; 1999, n = 1,573; and 2000, n = 1,185. The Y-axis shows effect sizes for the student's EHEEE 7-subject cumulative score, expressed as standard deviations. Red points and confidence intervals represent the relationship between soil iodine and test scores, while blue points and confidence intervals represent the placebo analysis for soil selenium. Coefficients for each birth year are shown relative to the reference year 1997. This placebo analysis is restricted to students residing in urban districts in the vicinity of Addis Ababa (Oromia Special Zone), where food supplies are expected to originate primarily from outside local agricultural systems and therefore should not reflect local soil iodine availability. Results for selenium serve as an additional placebo test because access to iodized salt would not be expected to alter the relationship between local selenium concentrations and academic achievement. All models control for sex, academic stream, and include fixed effects for test year, school, and district.

Figure S6. Event study of dose–response in exam scores to a 1 SD increase in local soil nutrients by year of birth, with conflict controls

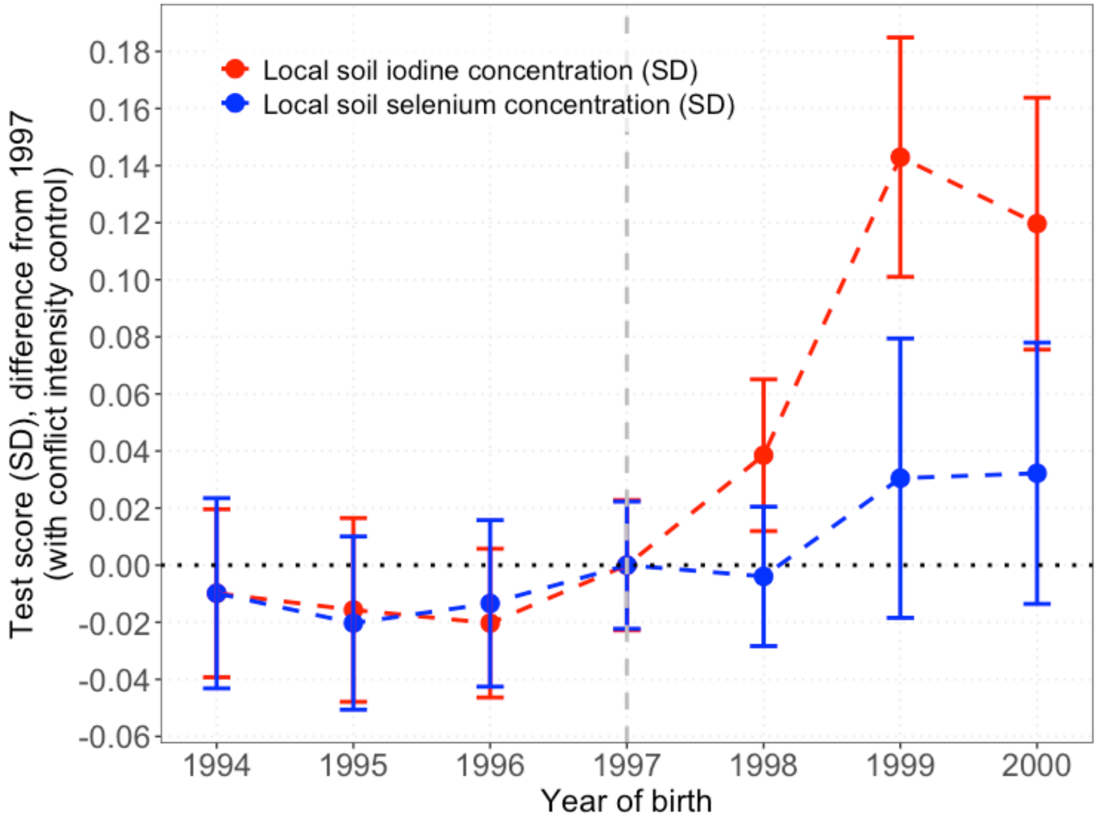

*Notes*: Data shown are estimated regression coefficients (points) with 95% confidence intervals calculated from heteroskedasticity-robust standard errors clustered at the district (woreda) level from the event-study dose-response analysis (Equation 3), re-estimated as a robustness check that additionally controls for district-year conflict exposure using ACLED measures of conflict events and fatalities. Statistical significance was assessed using two-sided tests, and no adjustment was made for multiple comparisons. The unit of analysis is the individual student exam record; no technical or biological replicates were used. The Y-axis shows effect sizes for the student's EHEEE 7-subject cumulative score, expressed as standard deviations. Red points and confidence intervals represent the relationship between soil iodine and test scores, while blue points and confidence intervals represent the falsification analysis for soil selenium. Coefficients for each birth year are shown relative to the reference year 1997, capturing differences in test scores associated with a one standard deviation higher level of local soil iodine across birth cohorts surrounding the 1998 disruption. The estimation sample is restricted to students residing in rural districts of the three study regions. All models control for sex, academic stream, district-year conflict exposure, and include fixed effects for test year and district. Details on the construction of the conflict measures are provided in the Supplementary Notes.

Figure S7. Event Study of Dose-Response in Child Health to 1 SD Higher Local Soil Iodine by Year of Birth

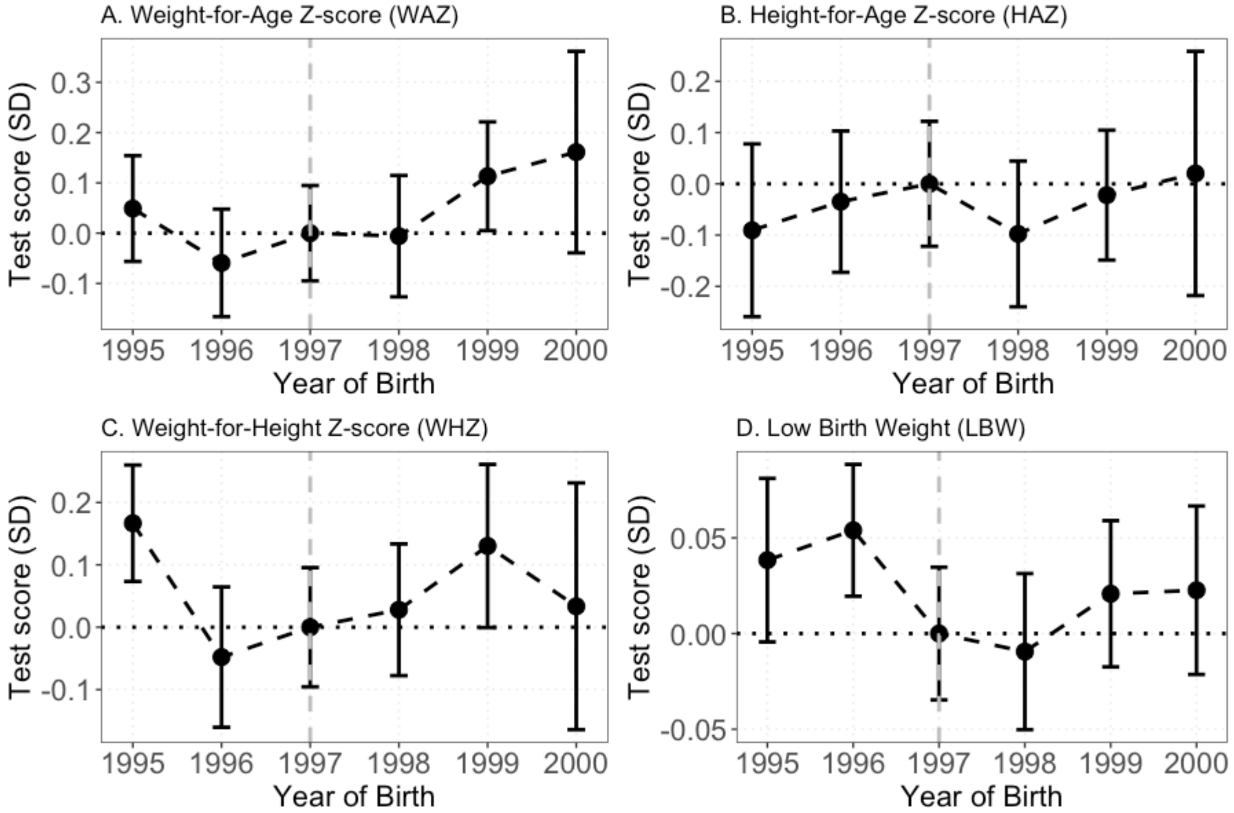

*Notes*: Data shown are estimated regression coefficients (points) with 95% confidence intervals calculated from heteroskedasticity-robust standard errors clustered at the woreda level from an event-study dose-response analysis. Statistical significance was assessed using two-sided tests, and no adjustment was made for multiple comparisons. The unit of analysis is the individual child; no technical or biological replicates were used. The Y-axis represents the estimated effect of a one standard deviation higher local soil iodine concentration on child health outcomes across birth cohorts from 1995 to 2000. Panels show weight-for-age Z-score (WAZ; A), height-for-age Z-score (HAZ; B), weight-for-height Z-score (WHZ; C), and low birth weight (LBW; D). Coefficients for each birth year are shown relative to the reference cohort born in 1997, the last full birth cohort before the 1998 war-induced disruption of iodized salt supplies. Models were estimated using survey-weighted linear regressions and control for child sex, household and maternal characteristics, and include survey-year and woreda fixed effects. The analysis is restricted to singleton births in rural districts, where children are more likely to rely on locally sourced foods.

Table S1. Summary statistics for selected variables from the Demographic and Health Surveys (DHS) and student exams (EHEEE)

| | **Full Sample** | | **1-Year Birth Window (Born 1997 or 1998)** | | **2-Years Birth Window (Born 1996-99)** | | **3-Years Birth Window (Born 1995-2000)** | |
|---|---|---|---|---|---|---|---|---|
| **Panel A: Child health (DHS)** | **N** | **Mean (SD)** | **N** | **Mean (SD)** | **N** | **Mean (SD)** | **N** | **Mean (SD)** |
| Time since birth (years) | 8,185 | 2.47 (1.43) | 1,787 | 2.26 (0.58) | 3,604 | 2.28 (1.15) | 4,769 | 2.64 (1.47) |
| Female (%) | 8,185 | 48.88 | 1,787 | 48.96 | 3,604 | 49.08 | 4,769 | 49.55 |
| Still alive (%) | 8,185 | 90.14 | 1,787 | 88.98 | 3,604 | 88.54 | 4,769 | 88.32 |
| Low birthweight (%) | 8,185 | 32.50 | 1,778 | 35.66 | 3,590 | 34.93 | 4,751 | 34.12 |
| Height-for-age z-score (HAZ) | 5,213 | -2.07 (1.71) | 1,502 | -2.55 (1.48) | 3,027 | -2.24 (1.59) | 3,808 | -2.18 (1.62) |
| Weight-for-height z-score (WHZ) | 5,214 | -0.60 (1.35) | 1,503 | -0.69 (1.20) | 3,023 | -0.69 (1.26) | 3,803 | -0.65 (1.28) |
| Weight-for-age z-score (WAZ) | 5,369 | -1.66 (1.34) | 1,531 | -1.93 (1.25) | 3,079 | -1.80 (1.26) | 3,887 | -1.75 (1.28) |
| Maternal age (years) | 6,270 | 29.33 (7.18) | 1,787 | 29.13 (7.10) | 3,604 | 29.21 (7.22) | 4,585 | 29.44 (7.21) |
| Maternal BMI ($kg/cm^2$) | 6,241 | 20.46 (6.19) | 1,776 | 20.15 (4.33) | 3,581 | 20.15 (4.52) | 4,556 | 20.25 (5.05) |
| Maternal edu. (>=primary, %) | 8,185 | 15.04 | 1,787 | 11.25 | 3,604 | 11.38 | 4,769 | 12.08 |
| Paternal edu. (>=primary, %) | 8,126 | 31.16 | 1,776 | 26.97 | 3,584 | 26.79 | 4,741 | 27.21 |
| Household wealth index | 8,184 | -0.04 (2.77) | 1,787 | 0.12 (3.41) | 3,603 | 0.12 (3.41) | 4,768 | 0.11 (3.34) |
| **Panel B: Exam scores (EHEEE)** | **N** | **Mean (SD)** | **N** | **Mean (SD)** | **N** | **Mean (SD)** | **N** | **Mean (SD)** |
| Age at test (years) | 1,080,685 | 19.46 (1.24) | 232,526 | 19.47 (1.03) | 452,136 | 19.45 (1.03) | 603,569 | 19.39 (1.08) |
| Female (%) | 1,080,685 | 39.82 | 232,526 | 44.67 | 452,136 | 45.31 | 603,569 | 45.6 |
| Natural Science track (%) | 1,080,685 | 62.28 | 232,526 | 62.14 | 452,136 | 62.33 | 603,569 | 63.09 |
| Aptitude score (SD) | 1,080,685 | -0.02 (0.97) | 232,504 | 0.02 (0.95) | 452,081 | 0.01 (0.95) | 603,483 | 0.00 (0.96) |
| English score (SD) | 1,080,685 | -0.05 (0.96) | 232,515 | -0.04 (0.94) | 452,114 | -0.03 (0.94) | 603,522 | -0.03 (0.95) |
| Civics score (SD) | 1,072,717 | 0.01 (0.96) | 232,494 | 0.05 (0.95) | 452,059 | 0.06 (0.97) | 603,445 | 0.05 (0.97) |
| Natural Math score (SD) | 673,098 | -0.03 (0.97) | 144,490 | 0.01 (0.97) | 281,813 | 0.00 (0.97) | 380,816 | -0.00 (0.97) |
| Biology score (SD) | 604,916 | 0.02 (0.98) | 144,470 | 0.05 (0.96) | 281,774 | 0.06 (0.98) | 380,749 | 0.05 (0.98) |
| Chemistry score (SD) | 604,925 | 0.02 (0.98) | 144,474 | 0.06 (0.97) | 281,775 | 0.07 (0.98) | 380,751 | 0.05 (0.98) |
| Physics score (SD) | 604,953 | -0.02 (0.97) | 144,476 | 0.02 (0.97) | 281,782 | 0.01 (0.97) | 380,763 | 0.00 (0.97) |
| Social Math score (SD) | 407,159 | 0.01 (0.99) | 88,007 | 0.09 (0.98) | 170,138 | 0.03 (0.98) | 222,488 | 0.03 (0.98) |
| Geography score (SD) | 323,922 | 0.05 (0.99) | 88,017 | 0.08 (0.98) | 170,278 | 0.078 (0.98) | 222,689 | 0.07 (0.98) |

| | | | | | | | | |
|---|---|---|---|---|---|---|---|---|
| History score (SD) | 323,922 | 0.05 (1.00) | 88,010 | 0.08 (1.00) | 170,265 | 0.09 (1.01) | 222,675 | 0.07 (1.01) |
| Economics score (SD) | 323,929 | 0.05 (0.99) | 88,017 | 0.08 (0.98) | 170,275 | 0.09 (0.99) | 222,683 | 0.07 (0.99) |
| All-Subjects average (SD) | 1,080,685 | -0.00 (0.97) | 232,526 | 0.05 (0.94) | 452,136 | 0.05 (0.95) | 603,569 | 0.04 (0.95) |
| **Panel C: District-Level local nutrient levels** | **N** | **Mean (SD)** | **Min** | **Max** | | | | |
| Organic soil iodine (mg/kg) | 385 | 10.18 (4.17) | 1.19 | 23.05 | | | | |
| Soluble soil selenium (µg/kg) | 385 | 1.85 (1.10) | 0.009 | 5.15 | | | | |
| Crop iodine (µg/kg) | 385 | 0.025 (0.027) | 0 | 0.22 | | | | |

*Notes*: This table provides summary statistics for variables used in our analysis, derived from the estimation sample outlined in Figures S10 and S11. **Panel A** aggregates data from two waves of the Demographic and Health Surveys (DHSs) conducted in 2000 and 2005. **Panel B** focuses on academic achievement, specifically students' performance on the EHEEE. Anthropometric indices measure deviations in a child's height or weight from the median of the reference population, based on WHO’s 2006 growth standards. Test scores are normalized into z-scores by centering on the mean and scaling by the standard deviation of students born in the same year, took the exam in the same year, and resided in the same administrative region. Data sources include the DHS, accessible upon registration at DHS Program, and academic achievement data provided by Ethiopia’s Ministry of Education. **Panel C** is our original data on soil nutrients and crop iodine.

Table S2. Summary statistics for selected variables from the Ethiopian National Micronutrient Survey (ENMS) of 2015/16

| Cohort | Variables | Panel A: All ENMS | | Panel B: ENMS with Crop Iodine Data | |
|---|---|---|---|---|---|
| | | N | Mean (SD)/ Median (p25, p75) | N | Mean (SD)/ Median (p25, p75) |
| School Aged Children (SAC) | Female (%) | 1,190 | 53.53 | 540 | 53.7 |
| | Age (Years) | 1,190 | 9.20 (2.72) | 540 | 9.34 (2.68) |
| | Median Urinary Iodine (µg/L) | 1,039 | 108 (66, 220) | 509 | 105 (65, 207) |
| | Goiter (%) | 1,190 | 5.38 | 540 | 3.89 |
| | Vitamin A Deficient (%) | 1,183 | 17.92 | 536 | 18.28 |
| | Vitamin B12 Deficient (%) | 932 | 10.41 | 400 | 11 |
| | Anemia (%) | 1,040 | 26.06 | 507 | 28.99 |
| Women of Reproductive Age (WRA) | Age (Years) | 1,195 | 27.94 (8.62) | 515 | 28.43 (8.77) |
| | Median Urinary Iodine (µg/L) | 1,013 | 97 (57, 178) | 436 | 86 (54, 166) |
| | Goiter (%) | 1,188 | 11.45 | 514 | 14.2 |
| | Vitamin A Deficient (%) | 1,174 | 4.26 | 505 | 4.16 |
| | Vitamin B12 Deficient (%) | 1,150 | 18.35 | 491 | 13.85 |
| | Anemia (%) | 1,195 | 19.16 | 515 | 15.73 |

*Notes*: This table presents summary statistics for selected variables from the 2015/16 Ethiopian National Micronutrient Survey (ENMS), shown separately for school-aged children (SAC) and women of reproductive age (WRA). Panel A reports statistics for the full ENMS sample, while Panel B restricts the sample to respondents residing in the three study regions (Tigray, Amhara, and Oromia) for whom crop iodine data are available. Continuous variables are reported as Mean (SD) or, for urinary iodine concentration, as Median (p25, p75), where p25 and p75 denote the 25th and 75th percentiles, respectively. Binary variables are reported as percentages. Urinary iodine concentrations are expressed in µg/L. The ENMS data were collected by the Ethiopian Public Health Institute (EPHI).

Table S3. Determinants of iodine concentration in grain samples from Amhara, Oromia and Tigray regions of Ethiopia

| | (1) | (2) | (3) | (4) | (5) | (6) | (7) | (8) |
|---|---|---|---|---|---|---|---|---|
| ln (Soil Organic Iodine) | 0.07*** | 0.12*** | | | | | | |
| SE | (0.025) | (0.029) | | | | | | |
| 95% CI | (0.021, 0.119) | (0.063, 0.177) | | | | | | |
| P value | 0.005 | 0.000 | | | | | | |
| ln (Soil Soluble Iodine) | | | 0.08*** | 0.10*** | | | | |
| SE | | | (0.031) | (0.032) | | | | |
| 95% CI | | | (0.019, 0.141) | (0.037, 0.163) | | | | |
| P value | | | 0.01 | 0.002 | | | | |
| ln (Soil Adsorbed Iodine) | | | | | -0.05*** | -0.03 | | |
| SE | | | | | (0.020) | (0.024) | | |
| 95% CI | | | | | (-0.089, -0.011) | (-0.077, 0.017) | | |
| P value | | | | | 0.012 | 0.211 | | |
| ln (Organic + Soluble Iodine) | | | | | | | 0.07*** | 0.12*** |
| SE | | | | | | | (0.026) | (0.029) |
| 95% CI | | | | | | | (0.019, 0.121) | (0.063, 0.177) |
| P value | | | | | | | 0.007 | 0.000 |
| ln (Altitude) | | -1.05*** | | -1.07*** | | -0.97*** | | -1.05*** |
| SE | | (0.304) | | (0.304) | | (0.308) | | (0.304) |
| 95% CI | | (-1.646, -0.454) | | (-1.666, -0.474) | | (-1.574, -0.366) | | (-1.646, -0.454) |
| P value | | 0.001 | | 0.000 | | 0.002 | | 0.001 |
| ln (Avg. Annual Temperature) | | -1.30*** | | -1.29*** | | -1.14** | | -1.30*** |
| SE | | (0.483) | | (0.484) | | (0.488) | | (0.483) |
| 95% CI | | (-2.247, -0.353) | | (-2.239, -0.341) | | (-2.096, -0.184) | | (-2.247, -0.353) |
| P value | | 0.007 | | 0.008 | | 0.019 | | 0.007 |
| ln (Avg. Annual Precipitation) | | -0.24*** | | -0.23*** | | -0.23** | | -0.24*** |
| SE | | (0.087) | | (0.088) | | (0.091) | | (0.087) |
| 95% CI | | (-0.411, -0.069) | | (-0.402, -0.058) | | (-0.408, -0.052) | | (-0.411, -0.069) |
| P value | | 0.006 | | 0.009 | | 0.011 | | 0.006 |
| Slope (degrees) | | 0.02*** | | 0.02*** | | 0.02*** | | 0.02*** |

| | | | | | | | | |
|---|---|---|---|---|---|---|---|---|
| SE | | (0.005) | | (0.005) | | (0.005) | | (0.005) |
| 95% CI | | (0.01, 0.03) | | (0.01, 0.03) | | (0.01, 0.03) | | (0.01, 0.03) |
| P value | | 0.000 | | 0.000 | | 0.000 | | 0.000 |
| ln (Distance to Nearest Coast) | | -0.12* | | -0.09 | | -0.07 | | -0.12* |
| SE | | (0.070) | | (0.069) | | (0.069) | | (0.070) |
| 95% CI | | (-0.257, 0.017) | | (-0.225, 0.045) | | (-0.205, 0.065) | | (-0.257, 0.017) |
| P value | | 0.086 | | 0.192 | | 0.31 | | 0.086 |
| ln (EVI) | | -0.00 | | 0.06 | | 0.17 | | -0.00 |
| SE | | (0.107) | | (0.105) | | (0.105) | | (0.107) |
| 95% CI | | (-0.21, 0.21) | | (-0.146, 0.266) | | (-0.036, 0.376) | | (-0.21, 0.21) |
| P value | | 0.99 | | 0.568 | | 0.105 | | 0.99 |
| N | 1,318 | 1,318 | 1,316 | 1,318 | 1,316 | 1,316 | 1,317 | 1,317 |
| R2 | 0.27 | 0.31 | 0.27 | 0.30 | 0.27 | 0.29 | 0.27 | 0.31 |

*Notes*: This table reports OLS estimates where the dependent variable is the natural logarithm of iodine concentration measured in staple grain samples, and the key independent variables are soil iodine fractions. The sample pools observations across ten staple grains (teff, wheat, maize, sorghum, barley, millet, rice, faba bean, chickpea, and lentil); crop-type fixed effects are included in all specifications. Each of the eight columns corresponds to a different model specification, varying the soil iodine measure (organic, soluble, adsorbed, or combined organic + soluble) and whether environmental covariates are included. Extended models additionally control for altitude, long-run climate (average annual temperature and precipitation), slope, distance to the coast, and vegetation index (EVI). Robust standard errors are shown in parentheses; 95% confidence intervals and exact p-values are reported below each estimate. Significance levels are denoted as *** $p < 0.01$, ** $p < 0.05$, * $p < 0.1$.

Table S4. Association of urinary iodine level (log) in children with their district average grain iodine concentration (log)

| | School Aged Children | | |
|---|---|---|---|
| | (1) | (2) | (3) |
| ln (Avg. Crop Iodine) | 1.17*** | 1.15*** | 1.07*** |
| SE | (0.408) | (0.379) | (0.328) |
| 95% CI | (0.37, 1.97) | (0.407, 1.893) | (0.427, 1.713) |
| P value | 0.004 | 0.002 | 0.001 |
| Indicator whether household uses adequately iodized salt | | | 0.34*** |
| SE | | | (0.119) |
| 95% CI | | | (0.107, 0.573) |
| P value | | | 0.004 |
| Controls | No | Yes | Yes |
| N | 509 | 499 | 474 |
| $R^2$ | 0.07 | 0.13 | 0.14 |

*Notes*: This table reports associations between individual urinary iodine concentration (UIC), measured from a single spot urine sample and log-transformed (µg/L), and the natural logarithm of average district-level iodine concentration in staple grains among school-aged children in rural districts of Amhara, Oromia, and Tigray[1]. Consistent with WHO guidance, UIC is interpreted only at the population level and is not used to classify individual iodine status. District-level grain iodine reflects naturally occurring iodine availability in local food systems and is constructed using the conditional simulation approach described in Supplementary Note A.3.B, aggregating iodine concentrations from maize, wheat, and teff[2]. Grain samples were collected during the 2017 and 2018 harvest seasons[2]. Model (1) includes no additional covariates; Model (2) adjusts for age (level and squared), sex, C-reactive protein (included as a general health covariate to control for infection burden), and household wealth; Model (3) further controls for household use of adequately iodized salt (>15 mg/kg). Urinary iodine data are drawn from the 2015/16 Ethiopian National Micronutrient Survey and do not overlap temporally with the 1998 iodized salt disruption period. These results are presented as supportive, suggestive evidence of the soil–grain–human iodine pathway motivating the exposure proxy used in the main analysis, rather than as causal estimates. Robust standard errors clustered at the enumeration area level are shown in parentheses; significance levels are *** p < 0.01, ** p < 0.05, * p < 0.1.

Table S5. Falsification test of dose response in exam scores to local soil selenium for children born after loss of access to iodized salt

| | Rural Districts (falsification test, due to lack of known impact on cognition) | | | Urban Districts around Addis Ababa (falsification test, due to use of food from elsewhere) | | |
|---|---|---|---|---|---|---|
| | Born 1998 vs 1997 | Born 1998-99 vs 1996-97 | Born 1998-2000 vs 1995-97 | Born 1998 vs 1997 | Born 1998-99 vs 1996-97 | Born 1998-2000 vs 1995-97 |
| Post | -0.023*** | -0.032*** | -0.036*** | -0.544 | -0.734 | -0.749 |
| SE | (0.008) | (0.01) | (0.011) | (0.348) | (0.748) | (0.869) |
| 95% CI | (-0.039, -0.006) | (-0.052, -0.011) | (-0.058, -0.013) | (-1.225, 0.138) | (-2.2, 0.731) | (-2.452, 0.953) |
| P value | (0.011) | 0.002 | 0.002 | 0.152 | 0.352 | -0.475 |
| Post × Local Selenium | 0.003 | 0.023 | 0.033* | -0.332 | -0.459 | -0.475 |
| SE | (0.011) | (0.017) | (0.019) | (0.221) | (0.474) | (0.554) |
| 95% CI | (-0.018, 0.023) | (-0.011, 0.056) | (-0.004, 0.071) | (-0.766, 0.102) | (-1.389, 0.47) | (-1.56, 0.611) |
| P value | 0.786 | 0.184 | 0.084 | 0.168 | 0.358 | 0.411 |
| Pre-war Avg. Score | 361.61 | 360.014 | 356.407 | 340.954 | 336.749 | 333.973 |
| SD | 63.928 | 63.928 | 63.928 | 76.82 | 76.82 | 76.82 |
| N | 232,393 | 451,877 | 603,208 | 5,171 | 9,794 | 13,735 |
| $R^2$ | 0.28 | 0.264 | 0.26 | 0.434 | 0.398 | 0.381 |

*Notes*: Data shown are estimated coefficients from the dose-response DID model described in Equation (2), using local selenium as a placebo variable in a falsification test for artifactual results due to model specification. The local selenium variable is constructed similarly as the iodine test in our main results, as z score of district-level average soil concentration. The dependent variable for all columns is the child's 3-subject cumulative score in English, Aptitude and Civics from the EHEEE, expressed as z scores. The bottom row of the table shows the mean and standard deviation of their raw scores to show magnitudes. Results for urban districts surrounding Addis Ababa (Oromia Special Zone) are also a placebo test, because food supplies to the city bring grain from elsewhere. Each column refers to cohorts born 1, 2 or 3 years after or before the loss of iodized salt in 1998 due to closure of the Ethiopia-Eritrea border, where the indicator variable 'Post' equals 1 for children born in or after calendar year 1998, and 0 otherwise. All models adjust for sex, academic stream, and include test year, school and district fixed effects. Standard errors, robust and clustered at the district level, are shown in parentheses. Significance levels are ***p<0.01, **p<0.05, *p<0.1.

Table S6. Robustness of the dose–response relationship between local soil iodine and secondary-school exam scores to conflict exposure

| | Born 1998 vs 1997 | Born 1998-99 vs 1996-97 | Born 1998-2000 vs 1995-97 |
|---|---|---|---|
| Post | -0.023*** | -0.037*** | -0.042*** |
| SE | (0.009) | (0.013) | (0.015) |
| 95% CI | (-0.042, -0.005) | (-0.063, -0.010) | (-0.071, -0.012) |
| P value | (0.013) | 0.006 | 0.006 |
| Post × Local Iodine | 0.024** | 0.080*** | 0.089*** |
| SE | (0.011) | (0.015) | (0.018) |
| 95% CI | (0.002, 0.046) | (0.051, 0.109) | (0.054, 0.123) |
| P value | 0.030 | 0.000 | 0.000 |
| Pre-war Avg. Score | 362.00 | 360.014 | 356.407 |
| SD | 64.287 | 63.928 | 64.287 |
| N | 232,393 | 451,877 | 603,208 |
| $R^2$ | 0.281 | 0.265 | 0.262 |

*Notes*: Data shown are estimated coefficients from the dose–response difference-in-differences model described in Equation (2), re-estimated as a robustness check to Table 1 for rural districts using iodine exposure. In addition to the baseline controls, these specifications further adjust for district-year conflict exposure, measured using Armed Conflict Location and Event Data (ACLED) indicators for total conflict events and conflict-related fatalities. The dependent variable in all columns is the child's cumulative score in English, Aptitude, and Civics from the Ethiopian Higher Education Entrance Examination (EHEEE), standardized as z-scores. The bottom rows report the mean and standard deviation of raw test scores to aid interpretation of magnitudes. Each column corresponds to cohorts born within 1-, 2-, or 3-year windows around the 1998 loss of iodized salt following the Ethiopia–Eritrea border closure; the indicator variable Post equals 1 for children born in or after 1998 and 0 otherwise. All models control for sex and academic stream, and include district, school, and test-year fixed effects. Standard errors are robust and clustered at the district level, shown in parentheses. Significance levels are ***$p < 0.01$, **$p < 0.05$, *$p < 0.1$.

Table S7. Assessing potential nonlinearity in the dose–response of secondary-school exam scores to local soil iodine

| | Born 1998 vs 1997 | Born 1998-99 vs 1996-97 | Born 1998-2000 vs 1995-97 |
|---|---|---|---|
| Post | -0.015 | -0.048** | -0.064** |
| SE | (0.005) | (0.022) | (0.025) |
| 95% CI | (-0.043, 0.014) | (-0.092, -0.005) | (-0.112, -0.015) |
| P value | (0.323) | 0.029 | 0.010 |
| Post × Local Iodine | 0.028** | 0.077*** | 0.083*** |
| SE | (0.011) | (0.015) | (0.018) |
| 95% CI | (0.007, 0.049) | (0.046, 0.107) | (0.048, 0.118) |
| P value | 0.011 | 0.000 | 0.000 |
| Post × Local Iodine$^2$ | -0.01 | 0.012 | 0.022 |
| SE | (0.011) | (0.015) | (0.018) |
| 95% CI | (-0.031, 0.011) | (-0.018, 0.042) | (-0.013, 0.056) |
| P value | 0.354 | 0.442 | 0.213 |
| Pre-war Avg. Score | 362.00 | 360.014 | 356.407 |
| SD | 64.287 | 63.928 | 64.287 |
| N | 232,393 | 451,877 | 603,208 |
| $R^2$ | 0.281 | 0.265 | 0.262 |

*Notes*: Estimates from dose–response difference-in-differences models that extend Equation (2) by allowing for curvature in iodine exposure through inclusion of a quadratic interaction between post-disruption birth cohorts and district-level soil iodine. The dependent variable is the child's standardized cumulative score in English, Aptitude, and Civics from the Ethiopian Higher Education Entrance Examination (EHEEE). Columns correspond to 1-, 2-, and 3-year birth-cohort windows around the 1998 loss of iodized salt. All models control for sex and academic stream and include district, school, and test-year fixed effects. Pre-war means and standard deviations of raw scores are shown for scale. Standard errors are robust and clustered at the district level (in parentheses). Significance levels: ***p < 0.01, **p < 0.05, *p < 0.1.

Table S8. Upper-tail robustness check for the dose–response relationship between local soil iodine and secondary-school exam scores

| | Born 1998 vs 1997 | Born 1998-99 vs 1996-97 | Born 1998-2000 vs 1995-97 |
|---|---|---|---|
| Post | -0.021** | -0.035** | -0.043*** |
| SE | (0.010) | (0.014) | (0.016) |
| 95% CI | (-0.039, -0.020) | (-0.063, -0.008) | (-0.074, -0.011) |
| P value | (0.032) | 0.012 | 0.008 |
| Post × Local Iodine | 0.033*** | 0.083*** | 0.086*** |
| SE | (0.012) | (0.016) | (0.020) |
| 95% CI | (0.010, 0.056) | (0.051, 0.114) | (0.048, 0.125) |
| P value | 0.005 | 0.000 | 0.000 |
| Post × Top-Iodine-Decile | -0.078 | -0.028 | 0.024 |
| SE | (0.063) | (0.079) | (0.088) |
| 95% CI | (-0.201, 0.045) | (-0.182, 0.127) | (-0.149, 0.196) |
| P value | 0.213 | 0.726 | 0.789 |
| Pre-war Avg. Score | 362.00 | 360.014 | 356.407 |
| SD | 64.287 | 63.928 | 64.287 |
| N | 232,393 | 451,877 | 603,208 |
| $R^2$ | 0.281 | 0.265 | 0.262 |

*Notes*: Estimates are from dose–response difference-in-differences models that extend Equation (2) by interacting post-disruption birth cohorts with both continuous district-level soil iodine and an indicator for districts in the top decile of the iodine distribution. This specification tests whether marginal effects attenuate or reverse at high iodine levels. The dependent variable is the child's standardized cumulative score in English, Aptitude, and Civics from the Ethiopian Higher Education Entrance Examination (EHEEE). Columns correspond to 1-, 2-, and 3-year birth-cohort windows around the 1998 loss of access to iodized salt. All models control for sex and academic stream and include district, school, and test-year fixed effects. Pre-war means and standard deviations of raw scores are reported for scale. Standard errors are robust and clustered at the district level (in parentheses). Significance levels: ***p < 0.01, **p < 0.05, *p < 0.1.

Table S9. Bootstrap robustness check of dose-response DID estimates of exam scores to local soil iodine

| | Born 1998 vs 1997 | Born 1998-99 vs 1996-97 | Born 1998-2000 vs 1995-97 |
|---|---|---|---|
| Post | -0.024*** | -0.036*** | -0.041*** |
| SE | (0.009) | (0.013) | (0.015) |
| 95% CI | (-0.042, -0.005) | (-0.062, -0.010) | (-0.070, -0.011) |
| P value | (0.011) | 0.007 | 0.006 |
| Post × Local Iodine | 0.025** | 0.080*** | 0.088*** |
| SE | (0.012) | (0.016) | (0.017) |
| 95% CI | (0.001, 0.048) | (0.047, 0.108) | (0.054, 0.121) |
| P value | 0.037 | 0.000 | 0.000 |
| N | 232,393 | 451,877 | 603,208 |
| Bootstrap Replicates | 500 | 500 | 500 |

*Notes*: This table reports bootstrap-based estimates from the dose-response difference-in-differences model corresponding to Table 1. A nonparametric joint bootstrap with 500 replications was used, simultaneously resampling districts and individuals within districts to account for both sampling variability and uncertainty in interpolated soil iodine exposure. Reported values include the bootstrap mean estimate, bootstrap standard error, 95% percentile confidence interval, and bootstrap p-value. Each column corresponds to cohorts born within 1-, 2-, and 3-year windows before and after the 1998 loss of access to iodized salt. Post equals 1 for children born in or after 1998, and 0 otherwise. Soil Iodine is the standardized district-level soil iodine measure. All models adjust for sex and academic stream and include test year, school, and district fixed effects. The final rows report the estimation sample size and the number of successful bootstrap replications used. Significance levels: ***p < 0.01, **p < 0.05, *p < 0.1.

Table S10. Falsification test of dose-response in growth of children born in the years after loss of iodized salt across the three birth cohorts

| | (1) | (2) | (3) | (4) |
|---|---|---|---|---|
| **Panel A: Born 1998 vs 1997** | WAZ | HAZ | WHZ | Pr [< Avg. Birthweight] |
| Post | 0.02 | 0.34 | -0.2 | 0.01 |
| SE | (0.239) | (0.265) | (0.189) | (0.029) |
| 95% CI | (-0.448, 0.488) | (-0.179, 0.859) | (-0.57, 0.17) | (-0.047, 0.067) |
| P value | 0.933 | 0.199 | 0.29 | |
| Post × ln (Soil Iodine) | 0.02 | -0.06 | 0.01 | -0.01 |
| SE | (0.122) | (0.139) | (0.125) | (0.048) |
| 95% CI | (-0.219, 0.259) | (-0.332, 0.212) | (-0.235, 0.255) | (-0.104, 0.084) |
| P value | 0.87 | 0.666 | 0.936 | 0.835 |
| N | 1,520 | 1,492 | 1,493 | 1,764 |
| $R^2$ | 0.18 | 0.21 | 0.18 | 0.12 |
| | (1) | (2) | (3) | (4) |
| **Panel B: Born 1998-99 vs 1996-97** | WAZ | HAZ | WHZ | Pr [< Avg. Birthweight] |
| Post | 0.05 | -0.11 | 0.00 | 0.07*** |
| SE | (0.142) | (0.171) | (0.135) | (0.021) |
| 95% CI | (-0.228, 0.328) | (-0.445, 0.225) | (-0.265, 0.265) | (0.029, 0.111) |
| P value | 0.725 | 0.52 | 0.99 | 0.001 |
| Post × Soil Iodine | 0.06 | -0.03 | 0.08 | -0.02 |
| SE | (0.099) | (0.123) | (0.116) | (0.032) |
| 95% CI | (-0.134, 0.254) | (-0.271, 0.211) | (-0.147, 0.307) | (-0.083, 0.043) |
| P value | 0.544 | 0.807 | 0.49 | 0.532 |
| N | 3,059 | 3,008 | 3,003 | 3,563 |
| $R^2$ | 0.16 | 0.21 | 0.15 | 0.11 |
| | (1) | (2) | (3) | (4) |
| **Panel C: Born 1998-2000 vs 1995-97** | WAZ | HAZ | WHZ | Pr [< Avg. Birthweight] |
| Post | 0.08 | -0.1 | 0.04 | 0.06*** |
| SE | (0.096) | (0.12) | (0.098) | (0.017) |

| | | | | |
|---|---|---|---|---|
| 95% CI | (-0.108, 0.268) | (-0.335, 0.135) | (-0.152, 0.232) | (0.027, 0.093) |
| P value | 0.405 | 0.405 | 0.683 | 0.000 |
| Post × Soil Iodine) | 0.06 | -0.01 | 0.03 | -0.02 |
| SE | (0.088) | (0.113) | (0.102) | (0.027) |
| 95% CI | (-0.112, 0.232) | (-0.231, 0.211) | (-0.17, 0.23) | (-0.073, 0.033) |
| P value | 0.495 | 0.929 | 0.769 | 0.459 |
| N | 3,059 | 3,008 | 3,003 | 3,563 |
| $R^2$ | 0.19 | 0.23 | 0.13 | 0.9 |

*Notes*: This table presents coefficients from the dose-response DID model specified in Equation (2), using anthropometric and health data from two waves of the DHS (2000 and 2005). The table is divided into three panels, each representing results for different birth cohort comparisons: Panel A compares children born in 1998 versus 1997 (1-year birth window); Panel B compares those born in 1998–99 versus 1996–97 (2-year birth window); and Panel C compares children born in 1998–2000 versus 1995–97 (3-year birth window). Falsification test outcomes include z-scores for weight-for-age (WAZ), height-for-age (HAZ), and weight-for-height (WHZ) relative to the 2006 WHO reference population, as well as an indicator for whether the child's mother reported below-average birthweight (used due to the absence of measured birthweight data). Null results are expected for these outcomes, as no prior evidence or mechanisms link iodine to physical growth. For each panel, the variable Post indicates children born in the later cohort (e.g., 1998 in Panel A), while ln(Soil Iodine) is the natural log of district-level average soil organic iodine concentration. The interaction term Post × ln(Soil Iodine) measures the differential effect of iodine availability across cohorts. Column (4) controls for all covariates except child age, while Columns (1)–(3) include age (level, squared, and cubed), birth order, maternal and paternal education, household wealth index, and whether the child was born in a medical facility. All models control for district and survey-year fixed effects, which account for time-invariant district characteristics and survey-year shocks. Pr[< Avg. Birthweight] refers to the probability of being born below average birthweight. Robust standard errors (in parentheses) account for the DHS sampling design. Significance levels are indicated as *** p<0.01, ** p<0.05, * p<0.1.

Figure S8. Flow chart of sample size and exclusion criteria used for estimation of effects on academic achievement

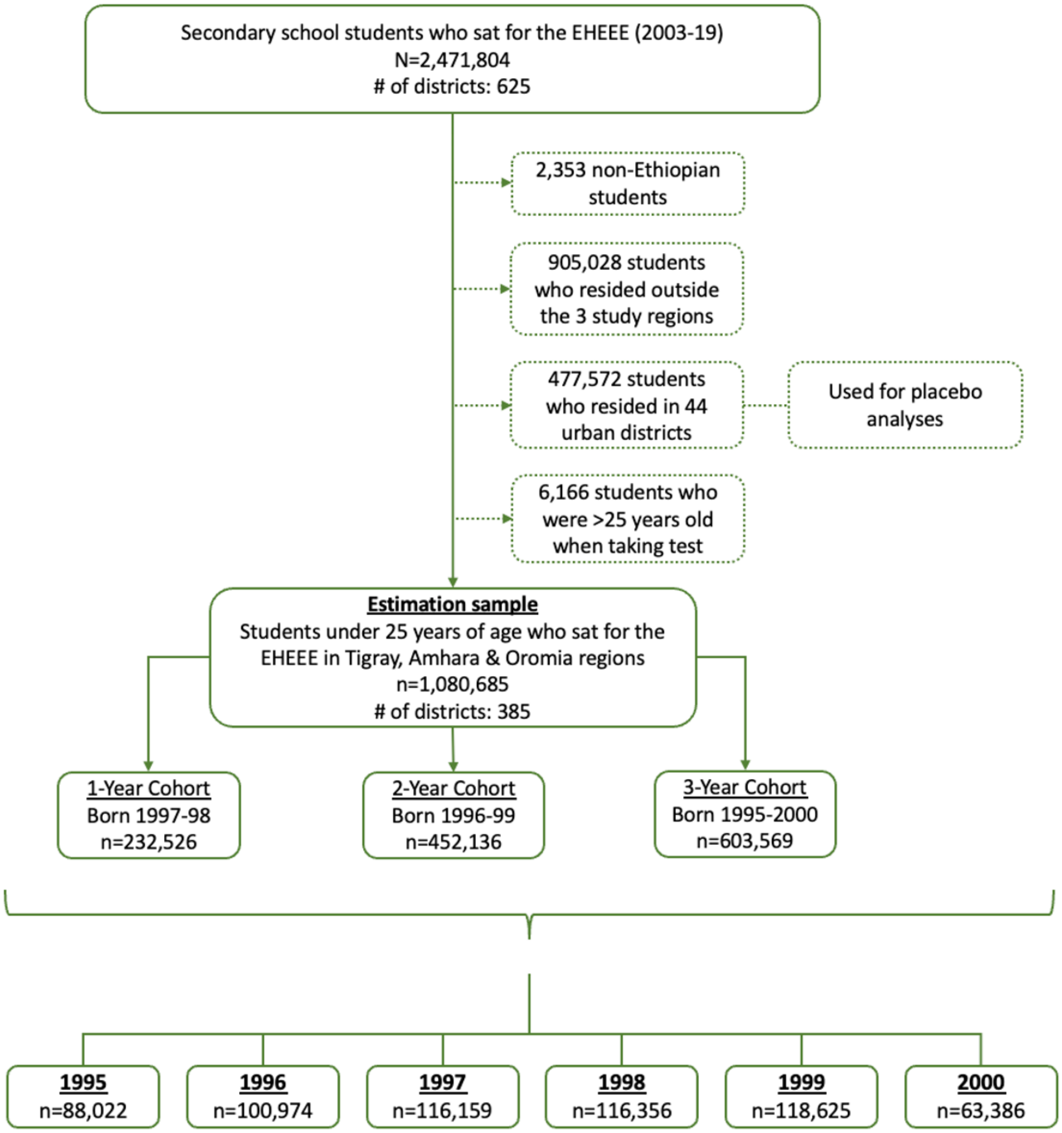

Figure S9. Flow chart of sample sizes and exclusion criteria used for estimation of effects on mortality and physical growth

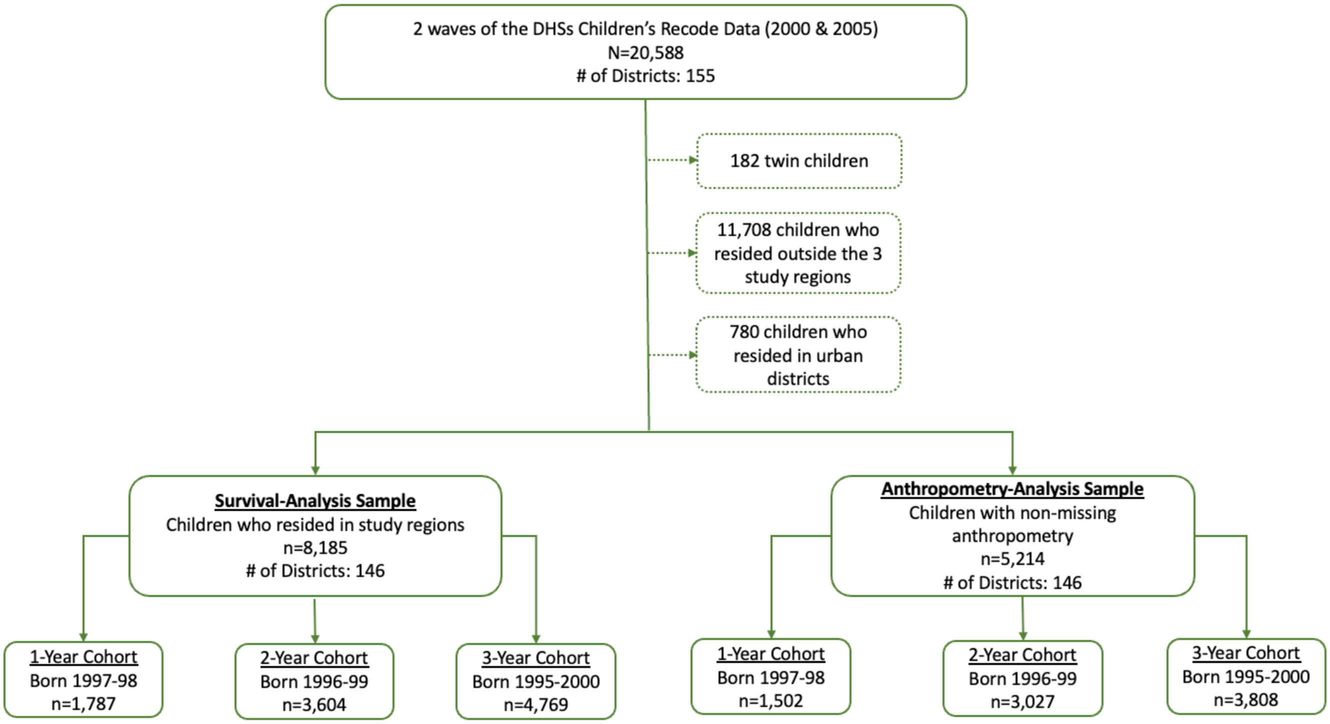

**Supplementary Notes**

**A1. Soil and grain sampling procedure**

The purpose of field sampling was to support spatial mapping, by point prediction and prediction of regional means for administrative units, including districts, nutrient concentration in crops and associated soils, using methods previously reported[1] based on a spatial multivariate linear mixed model (LMM) [3]. The sampling frame covered three major regions of Ethiopia (Amhara, Oromia and Tigray), based on a gridded map at 500-m resolution developed by the AfSIS project [4] using information from high-resolution satellite imagery and multiple covariates obtained from remote sensor data and digital elevation models [5]. From that map we retained all map pixels where the probability of agricultural land use was estimated to be at least 90 percent, and which fell within 2.5 km of a known road as recorded in OpenStreetMap [6]. This constituted the sample frame. A set of primary sample locations were then selected from the sampling frame using the *lcube* package from the *BalancedSampling* library of the R platform [7]. This package allows one to sample sites satisfying specified inclusion probabilities while ensuring spatial balance and spread. A subset of the primary sample sites was selected by spatially balanced sampling and earmarked for a second field sample site at a nearby location. These extra close-pair sites were included to support estimation of parameters of the spatial statistical model. In the original sample design, a set of 1825 primary sample locations were selected, with an additional 175 close-pair sites [2,8]. The final sample set consisted of 1128 primary locations and 224 close pair locations, 1352 sample sites in total, with loss of sample numbers where sites could not be accessed, where post-sampling quality checks cast doubt on the location of a sample (grain or soil), or where analytical results for iodine or selenium were rejected during quality control.

Field sampling was conducted by teams of trained experts [9]. First, the team located the sampling site and looked for the nearest field with a mature cereal crop within a 1-km radius and requested permission from the farmer to sample both the soil and the grain. Cereals were identified by the team as either barley, field pea, finger millet, maize, pearl millet, rice, sorghum, teff, triticale, or wheat. If teams did not observe a field with a standing mature cereal crop, they would ask local farmers to identify a nearby field from which they had recently harvested any of these cereals, and that field was sampled if permission was obtained to sample soil and the grain from that field, from a field stack or store according to circumstances. Soil samples were collected from a 100-$m^2$ (0.01-ha) circle as close as the middle of the selected field. Within that circle, five points were identified based on the orientation described in [9] . One soil sample was collected at each of the five points using a Dutch auger (150 mm length by 50 mm diameter). To reduce the risk of contamination, any plant material sticking to the auger was removed, and the five samples then emptied into a single bag. The combined soil sample was then matched with a crop sample from the same location, using either standing or recently harvested grain.

When a standing crop was present in the field it was sampled at the same five locations as the soil samples and a single aggregate grain sample was produced. Crops with heads (e.g., wheat, barley), were sampled by cutting heads with sterilized stainless-steel scissors. In the case of maize, one sample cob was harvested at each auguring position. All harvested crop heads and maize cobs were placed into pre-labeled paper sample bags. Once collected, the crop heads and maize cobs were manually threshed or shelled to separate the grains. This process was performed under clean conditions to minimize external contamination. More information on the field protocols is provided in prior publications [2].

**A2. Soil and grain sample preparation and laboratory analyses**

Details of the soil and grain sample preparation and analyses are described in prior publications [8]. Briefly, soil samples were oven-dried and sieved to pass through 2 mm. Dried grain samples were ground using a stainless-steel coffee grinder. The ground samples were transferred into clean, pre-labeled polyethylene bags and sealed tightly to prevent moisture uptake. The prepared soil (150 g) and grain samples (~20 g) were sent to the University of Nottingham and Rothamsted Research for a range of assays. For soil and grain iodine, extractions and analysis using inductively coupled plasma mass spectrometry (ICP-MS).

For soils, a three-step sequential extraction procedure adapted from prior studies [10] was used to estimate three fractions of iodine and selenium nominally identified as 1) "soluble" fraction in 0.01 M $KNO_3$, 2) "adsorbed fraction" in 0.016 M $KH_2PO_4$ and 3) "organically bound fraction" in 10% tetramethyl ammonium hydroxide (TMAH). A mass of air-dried soil sample (≈ 4.0 g), sieved to

<4 mm, was weighed into a polyethylene centrifuge tube. After adding 20 mL of 0.01 M $KNO_3$, the tubes were shaken for 2 hours on an end-over-end shaker, then centrifuged at 3500 rpm for 30 minutes. A volume of 9 mL of the supernatant solutions was filtered to <0.22 µm using PTFE syringe filters, into a 14 mL tube containing 1 mL of 10% TMAH. After removing the excess supernatant, the centrifuge tubes with wet soil pellets were weighed to account for carry over of 0.01 M $KNO_3$ extract. Then, 20 mL of 0.016 M $KH_2PO_4$ was added to the remaining soil pellet. The tubes were vortexed to disaggregate the soil pellet and then shaken for 1 hour on end-over-end shaker, then centrifuged at 3500 rpm for 30 minutes. A volume of 9 mL of the supernatant solutions was filtered to < 0.22 µm using PTFE syringe filters, into tubes containing 1 mL of 10% TMAH. After removing the excess supernatant, the centrifuge tubes with wet soil pellets were weighed again to account for carry over of 0.016 M $KH_2PO_4$ extract. Finally, 10 mL of 10% TMAH was added to the remaining soil pellet. The tubes were vortexed to disaggregate the soil pellet then loosely capped and incubated at a maximum of 70 $^0$C for ~ 16 hours. After incubation, the tubes were centrifuged at 3500 rpm for 30 minutes then 1 mL of the TMAH extracts were diluted with 9 mL of ultrapure MQ water to give a solution of 1% TMAH. Samples were analyzed using an inductively coupled plasma mass spectrometer (ICP-MS) equipped with triple quadrupole capability (iCAP-Q; Thermo Fisher Scientific, Bremen, Germany), operated in hydrogen cell mode. Rhenium ($^{187}$Re; 20 µg/ L) was used as internal standard to correct for instrumental drift.

The iodine composition of the grain samples was analyzed using inductively coupled plasma mass spectrometry (ICP-MS) as described previously [11]. A mass of milled grain sample (≈400 mg) was extracted in PTFE-TFM pressure-activated-venting vessels using 6 mL of 5% TMAH in a Multi-wave Pro Platform with 41HVT56 Rotor (Anton Paar GmbH, Graz, Austria) reaction system. Program settings were as follow: (I) 10 minutes ramp up to 140 °C, (II) holding at 140 °C for 20 minutes at a power of 1500 W, and (III) cooling to 55 °C over 15 minutes. After cooling, the extract was diluted to 30 mL with ultrapure water MQ-water, to give a solution of 1% TMAH, prior to analysis using ICP-MS equipped with triple quadrupole capability (iCAP-Q; Thermo Fisher Scientific, Bremen, Germany) operated in standard mode. Rhenium ($^{187}$Re; 5 µg/L) was used as internal standard to correct for instrumental drift. A certified reference material, hay powder BCR-129 (Institute for Reference Materials and Measurements, Geel, Belgium), was included to assess the accuracy of the digestion and analysis. The recovery for iodine in BCR hay powder was 89.6% ± 6.61%.

##### A3. Geospatial prediction of district-level soil and crop nutrient concentrations

###### A. Prediction (conditional simulation) of district-level mean soil nutrient concentration

An optimal method to upscale from the point observations to district-level average soil mineral nutrient concentrations, in the sense that it produces the best linear unbiased prediction from the data is block kriging [12] , and this was used on other variables collected in this project[1]. However, the soil iodine data are log-normally distributed, so the block kriging prediction for a sub-region (in this case, a district) cannot be simply back-transformed to the original scale of the measurement without the implausible assumption that the log-normality of mineral concentrations is invariant with the change of support from core to district [13]. For this reason, we used a conditional simulation approach. After exploratory analysis, we modeled the spatial pattern of soil organic iodine and soluble selenium concentrations as a step function of latitude, with a change in mean at 10 degrees. The residuals from the mean value north and south of this threshold were computed, and their summary statistics were examined. In both cases, the residuals showed marked positive skewness. The residuals were then transformed to normality using the Gaussian anamorphosis method, which is based on Hermite polynomials, using the anam.fit function from the *RGeostats* package for the R platform [14]. The number of polynomials was adjusted by trial and error to produce a smooth transformation. The variograms of the transformed residuals were then estimated using the standard estimator [15] and the robust alternatives described elsewhere [16]. The exponential variogram model was then fitted with these estimates as it is authorized for modeling the variogram on the sphere when coordinates are given as longitude and latitude [17]. Following this, the three models were assessed by cross-validation, and examination of the median standardized square prediction error [18] and the coverage probabilities of confidence intervals based on the kriging variance [19]. This provides a basis for confidence in simulation results generated from these models.

Each district was represented as a discretization on a square grid at intervals of 1,500 meters. A set of values of the transformed residual at each grid point was obtained by conditional simulation, conditioned on both the selected variogram model and the observed values at sample locations. The conditional simulation was done by the LU decomposition method [20]. A total of 10,000 realizations of the transformed residual variable over the discretization grid were generated. For each realization, the transformed residuals were back-transformed to the original scale of measurement, and added to the mean value for the original variable either north or south of 10 degrees latitude. These simulated organic iodine and soluble selenium values were then averaged over the grid nodes in each district to compute a conditional mean value. The empirical distribution of 10,000 simulated mean values for each district was then used to compute an empirical estimate of the district mean and quantiles of its conditional distribution.

#### B. Prediction of grain nutrient concentration

District-scale mean grain concentrations (maize, wheat, and teff) of iodine were obtained from the grain I data. As I concentrations in the grains were markedly non-normal, the same approach was used for soil I as described above. There was no need for a spatial function to account for a marked change in the mean, as for the soil data. The grain I concentrations were transformed to resemble values from a normal distribution by Gaussian anamorphosis, as described above. Variogram models were estimated, validated, and selected, and then used to generate realizations of the conditional distribution of the transformed variable at discretization nodes. The values for each realization were back-transformed, averaged separately over the nodes in each district, and these values were used over multiple realizations to obtain an empirical estimate of the conditional distribution of maize, wheat or teff I concentration for each district.

The resulting conditional mean values for each district were used as the district-level measures of organic soil iodine, soluble soil selenium, and crop iodine in the empirical analyses. To visualize this spatial variation, the predicted district-level mean concentrations for organic soil iodine and soluble soil selenium were joined to Ethiopian district boundary shapefiles using district identifiers and mapped in ArcGIS with graduated colour classes. This mapping workflow generated Figure 2 directly from the spatial boundary data and predicted district-level nutrient estimates; no previously published maps, map images, or third-party cartographic artwork were reproduced or adapted.

#### A4. Additional robustness analyses

##### A. Boostrap estimation incorporating soil-iodine prediction uncertianity

A potential robustness concern in our setting is that the district-level soil-iodine measures used in Equation (2) are not directly observed quantities but rather conditional simulations derived from a geostatistical prediction procedure (Section A3). These simulated district means incorporate uncertainty arising from spatial interpolation, the estimated variogram of transformed residuals, and the back-transformation from Gaussian anamorphosis. Treating these predicted iodine values as fixed regressors in the main specification may therefore understate total uncertainty if the dose–response estimates are sensitive to either geospatial prediction noise or sampling variability in the exam-score data.

To assess whether our estimates remain stable under these sources of uncertainty, we implement a bootstrap procedure that jointly perturbs both the environmental exposure and the individual-level outcomes. The procedure resamples districts with replacement, thereby repeatedly drawing from the empirical distribution of the district-level iodine simulations generated in Section A3. Within each selected district, exam takers are resampled with replacement, capturing sampling variability while preserving the district structure required by the quasi-experimental design. For each bootstrap replication, the full specification in Equation (2) is re-estimated using the same fixed effects as in the main analysis.

We conduct 500 bootstrap iterations, yielding an empirical distribution of the treatment and dose–response coefficients that reflects uncertainty from both the geospatial exposure measure and the exam-score data. Implementation in R follows this structure by reconstructing datasets through district-level and within-district resampling and re-estimating Equation (2) via felm(). The resulting percentile intervals provide a stringent robustness check, confirming that the estimated effects are stable once uncertainty in soil-iodine prediction and student-level sampling variability are jointly propagated through the model.

##### B. Assessing potential non-linearity in the dose-response relationship

Another potential robustness concern is that the relationship between iodine exposure and cognitive outcomes may not be strictly linear. Biological evidence suggests larger cognitive gains when populations move from deficiency toward sufficiency, potential plateaus thereafter, and risks at extremely high intake levels. Consistent with this concern, we already assess nonlinearity in the main analysis by replacing continuous district-level iodine with terciles of the soil-iodine distribution. To further evaluate whether the linear specification of Equation (2) adequately captures the exposure–response pattern, we implement two complementary extensions that allow for departures from linearity.

First, we extend Equation (2) to allow curvature by including a quadratic interaction in district-level iodine. Formally, we augment Equation (2) by adding a squared term in \text{Iodine}_d interacted with $Post_t$:

$$Y_{idt} = \alpha + \beta Post_t + \delta_1(Post_t \times Iodine_d) + \delta_2(Post_t \times Iodine_d^2) + \tau_d + \mu_s + X_{idt}\theta + \varepsilon_{idt}$$

This specification permits the marginal effect of iodine exposure following the disruption to vary flexibly across the distribution of \text{Iodine}_d, rather than imposing a constant linear gradient.

Second, to specifically probe potential departures from linearity at high exposure levels, we implement an upper-tail test by interacting \text{Post}_t with an indicator for districts in the top decile of the soil-iodine distribution. This specification allows the post-disruption iodine gradient to differ discretely for high-iodine districts, providing a targeted check for plateauing or adverse effects that may arise only at the upper end of the exposure range.

##### C. Controlling for spatial and temporal heterogeneity in conflict exposure

A further robustness concern is that the withdrawal of iodized salt in 1998 coincided with the Ethiopia–Eritrea conflict, which could have independently affected child health and educational outcomes through channels unrelated to iodine deficiency. Although the baseline specifications include district fixed effects, test-year fixed effects, and cohort timing, these controls do not fully absorb heterogeneity in conflict exposure across districts and years. If local conflict intensity varied systematically over time and space and correlated with underlying soil iodine levels, the estimated iodine effects could partly reflect residual war-related shocks rather than nutritional mechanisms.

To address this concern, we explicitly control for district-level conflict exposure using data from the Armed Conflict Location and Event Data Project (ACLED). Let d index districts and t index calendar years. For each district–year pair, we construct two complementary measures of conflict exposure. The first is the total number of recorded conflict events,

$$Events_{dt} = \sum_{e} 1\{e\ occured\ in\ district\ d\ in\ year\ t\}$$

where $e$ indexes individual ACLED conflict events. This measure captures the frequency of conflict-related incidents, regardless of their severity. The second measure captures conflict severity through total fatalities,

$$Fatalities_{dt} = \sum_{e \in (d,t)} fatalities_e$$

Since fatality counts are highly skewed, we use a log-transformed version i.e., $ConflictIntensity_{dt} = \log(1 + Fatalities_{dt})$, which reduces sensitivity to extreme values while preserving meaningful variation. District–year observations with no recorded conflict events are assigned zero exposure for both measures.

These conflict measures are merged into the student-level exam dataset using harmonized district identifiers and each child's birth year, capturing local conflict conditions during early-life exposure. We then augment the main dose–response specification in Equation (2) by including conflict exposure as an additional control. In the event-study framework, these controls enter alongside cohort-specific interactions between birth-year indicators and soil iodine, with coefficients normalized relative to the 1997 cohort.
